# A pan-cancer regulatory atlas of 6,983 GWAS variants prioritizes recurrent regulatory annotations and candidate programs at cancer risk loci

**DOI:** 10.64898/2026.05.16.26353369

**Authors:** Subhajit Dutta

**Affiliations:** Department of Biochemistry and Molecular Cell Biology, University Medical Center Hamburg-Eppendorf (UKE), Hamburg, Germany

## Abstract

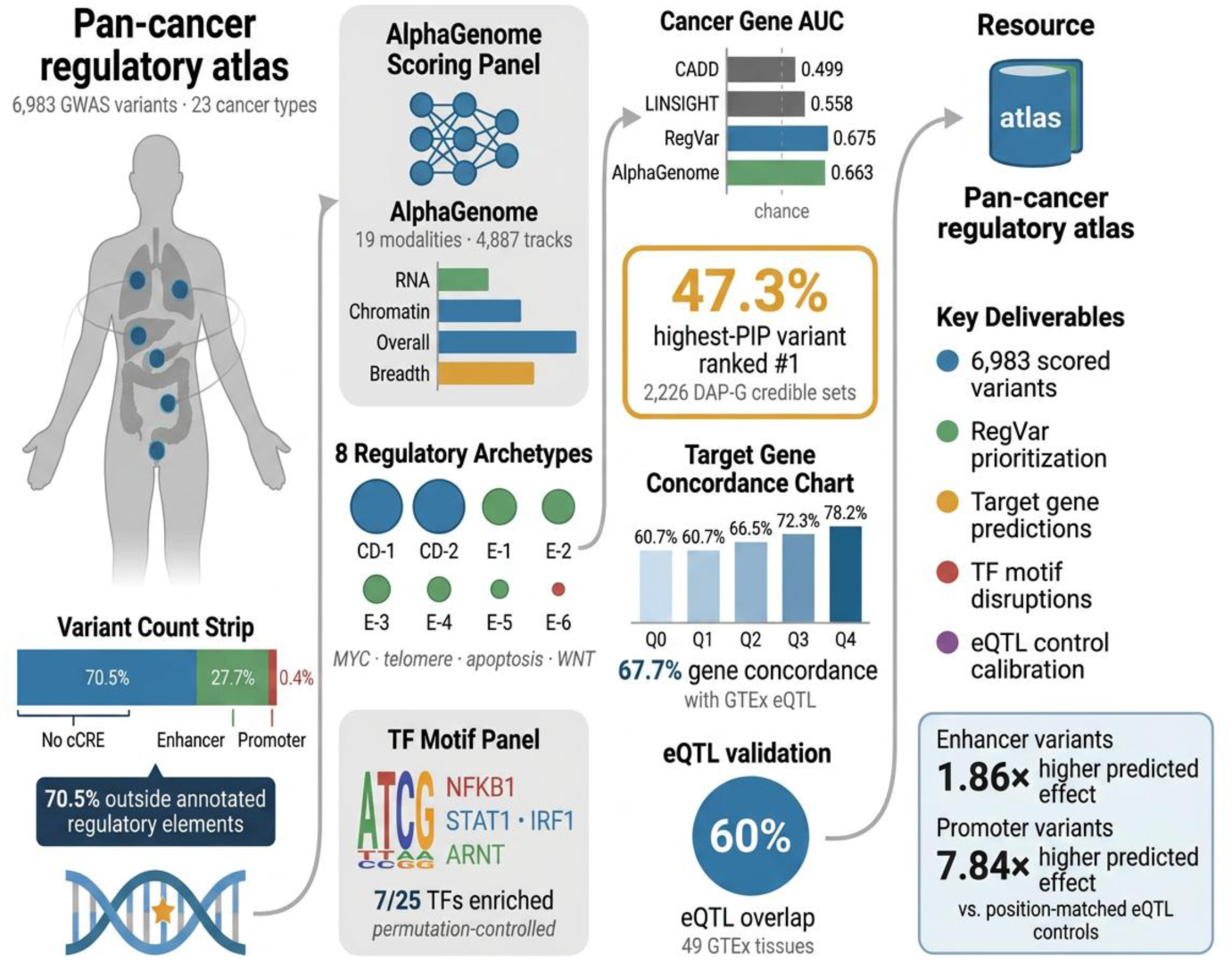

Genome-wide association studies have identified thousands of cancer risk variants in non-coding regions, yet their regulatory mechanisms remain largely uncharacterized. Here we present a regulatory annotation atlas of 6,983 genome-wide significant variants across 23 cancer types, scored using multimodal AlphaGenome predictions and integrated with ENCODE-4, Roadmap Epigenomics, and JASPAR 2024 annotations. Most variants (70.5%) fall outside annotated cis-regulatory elements; 27.7% overlap enhancers and 1.4% overlap promoters. Comparison with 6,626 position-matched eQTL control variants suggests that enhancer-classified variants carry 1.86-fold higher predicted effects (P = 10^−^⁹⁴) and promoter variants 7.84-fold (P = 2.5 × 10^−^¹⁹). A composite prioritization score (RegVar-basic, excluding GWAS-derived pleiotropy and TF disruption, AUC = 0.650; RegVar-full, AUC = 0.675) outperforms CADD (0.499) and LINSIGHT (0.558) in this cancer-gene discrimination benchmark. Within-locus ranking across 2,626 GTEx DAP-G eQTL credible sets shows that RegVar identifies the highest-posterior-probability variant in 47.3% of loci (P = 7.0 × 10^−^¹³), while CADD performs at chance. Predicted target genes show 67.7% concordance with GTEx eQTL assignments. Permutation-controlled motif analysis highlights NFKB1/NF-κB, STAT1, IRF1, and ARNT as exploratory permutation-enriched candidate transcription factors at cancer risk loci. This atlas provides a resource for interpreting non-coding cancer susceptibility variants. Because AlphaGenome uses expression-related training data, GTEx-based validations should be interpreted as partially orthogonal rather than fully independent.

## Introduction

Cancer susceptibility is substantially heritable, with twin studies estimating that genetic factors contribute 20–40% of risk for most common malignancies [1]. Over the past two decades, genome-wide association studies (GWAS) have identified thousands of risk variants, with extensively characterized breast cancer risk regions [2,3], hundreds of prostate cancer risk variants [4,5], and a substantial set of colorectal cancer susceptibility loci [6]. These discoveries have enabled the construction of polygenic risk scores with increasing clinical utility for risk stratification [7], yet a fundamental gap persists between statistical association and mechanistic understanding: approximately 93% of cancer GWAS variants reside in non-coding genomic regions where the rules governing regulatory function remain incompletely understood [8].

For a small number of landmark loci, the causal variant and its molecular mechanism have been established. Recurrent non-coding mutations at the TERT promoter create de novo ETS-family binding sites and are among the most frequent somatic non-coding alterations across cancer types [9]. Long-range enhancer regulation of MYC at the 8q24 pan-cancer risk locus [10] illustrates how distal regulatory elements can influence susceptibility across tissues of distinct developmental origin. However, for the vast majority of GWAS signals, the identity of the causal variant, the target gene, the cell type of action, and the molecular mechanism all remain unknown.

Multiple complementary computational strategies have been developed to address this interpretive gap. Statistical fine-mapping methods such as SuSiE [11] and FINEMAP [12] narrow GWAS signals to credible sets of candidate causal variants. Expression quantitative trait locus (eQTL) analyses and colocalization methods [13] connect variants to target genes in specific tissue contexts. Stratified LD score regression [14] quantifies the contribution of functional annotation categories to disease heritability at the genome-wide level. Integrative platforms including the Open Targets Genetics portal [15] and the ENCODE-4 candidate cis-regulatory element (cCRE) registry [16] provide systematic frameworks for variant-to-function annotation. Pan-cancer genetic analyses have characterized shared heritability across cancer types [17] and mapped pleiotropic susceptibility loci [18], but a comprehensive atlas linking individual variants to multimodal regulatory predictions across the full spectrum of human malignancies has been lacking.

The emergence of deep learning sequence-to-function models has created new opportunities for systematic variant effect prediction. Enformer [19] demonstrated that transformer-based architectures could predict gene expression and chromatin accessibility from 196-kb sequence windows. Most recently, AlphaGenome [20], developed by Google DeepMind, extended multimodal prediction to 19 distinct functional modalities—including RNA-seq across 667 cell types, chromatin accessibility, histone modifications, transcription factor binding, chromatin contacts, and splicing—from a 1,048,576-bp input window, representing the most comprehensive sequence-to-function model available to date.

Here we present a multimodal regulatory annotation atlas of 6,983 cancer GWAS variants across 23 cancer types, generated by systematic application of AlphaGenome variant effect prediction and integrated with the ENCODE-4 cCRE registry [16], Roadmap Epigenomics chromatin state annotations [21], JASPAR 2024 position weight matrix (PWM)-based transcription factor motif analysis with permutation controls [22], and extensive validation against GTEx v8 eQTL data [23], including within-locus fine-mapping comparison using DAP-G credible sets. We benchmark a composite prioritization score (RegVar) against CADD [24] and LINSIGHT [25], showing that these general-purpose variant prioritization scores perform at or near chance level for prioritizing regulatory annotations at non-coding cancer GWAS loci, whereas multimodal regulatory predictions achieve meaningful discrimination. We calibrate all results against 6,626 position-matched eQTL control variants scored using the identical pipeline, providing empirical assessment of score inflation due to multiple testing across tracks.

## Material and Methods

### Pan-cancer GWAS variant compilation

We extracted all cancer-associated variants reaching genome-wide significance (P < 5 × 10−8) from the NHGRI-EBI GWAS Catalog (release r2026-04-27; 1,099,366 total associations) [26]. Cancer associations were identified by mapping disease traits to Experimental Factor Ontology terms across 23 cancer type categories, supplemented by keyword matching. Treatment-response, survival, and toxicity associations were excluded to focus on germline susceptibility. After removing duplicate entries at identical genomic positions on GRCh38, this yielded 6,983 unique variants across 1,302 independent loci (defined by merging variants within 500-kb windows, a conservative distance threshold chosen to accommodate LD structure at cancer risk loci) spanning 23 cancer types. Pleiotropic variants, defined as those associated with two or more distinct cancer types, numbered 454 (6.5%) across 172 pleiotropic loci.

### AlphaGenome multimodal variant effect prediction

All 6,983 variants were scored using the AlphaGenome Python SDK via the public API [20]. For each variant, a 1,048,576-bp input sequence centered on the variant position was extracted from GRCh38. AlphaGenome returns predictions across 19 functional modalities: RNA-seq (667 cell types), ATAC-seq (167), DNase-seq (28), histone ChIP-seq for H3K27ac, H3K4me1, and H3K4me3 (1,617 combined tracks), transcription factor ChIP-seq (1,116 tracks), CAGE (546), PRO-CAP (12), Hi-C chromatin contacts, splice junctions, and splice site usage (734 tracks). For 1,678 variants (24.0%) with unspecified risk alleles in the GWAS Catalog, all three possible alternate alleles were scored and the maximum absolute effect retained. Four summary features were extracted per variant: maximum RNA-seq effect score, maximum chromatin effect score, number of modalities with detectable signal, and overall maximum effect score.

### Position-matched eQTL control variant generation and scoring

To provide empirical calibration for AlphaGenome score distributions, 6,626 position-matched eQTL control variants were generated by selecting, for each atlas variant, a control position from the GTEx v8 Whole Blood eQTL variant pool (1,277,232 unique positions) on the same chromosome within ±500 kb. Atlas positions were excluded from the control pool. All control variants were scored using the identical AlphaGenome pipeline—the same SDK version, the same max-of-3-alternate-alleles strategy, and the same feature extraction code—enabling direct distributional comparison free of processing artifacts. We note that the control pool consists of GTEx eQTL variants, which are themselves biologically active; this makes the comparison conservative, as does the ±500-kb proximity to atlas variants, which introduces partial LD sharing (Supplementary Table S7).

### Regulatory annotation and mechanism classification

Each variant was classified into regulatory categories by integrating AlphaGenome predictions with overlapping candidate cis-regulatory elements (cCREs) from the ENCODE-4 registry (1,063,878 human cCREs on GRCh38) [16]: promoter (PLS cCRE overlap; 1.4%), enhancer (pELS or dELS overlap; 27.7%), insulator (CTCF-only overlap; 0.4%), or putative distal regulatory (no annotated cCRE overlap; 70.5%). This last category encompasses variants that may act through unannotated enhancers or context-dependent regulatory effects not captured by current cCRE annotations. We note that this classification integrates cCRE annotations with AlphaGenome predictions by design; concordance between these sources reflects principled integration of complementary data rather than independent validation.

### Regulatory archetype discovery and stability assessment

A feature matrix was constructed from four quantitative features (maximum RNA-seq effect, maximum chromatin effect, maximum overall effect, number of modalities with signal) concatenated with one-hot-encoded mechanism class indicators, standardized to zero mean and unit variance. K-means clustering was performed for k = 3 to 11 with 20 random initializations per k, selecting optimal k = 8 by maximum mean silhouette coefficient (0.443). Archetype stability was assessed by 100 bootstrap resamples with computation of the adjusted Rand index (ARI = 0.949 ± 0.118). As a robustness check, enriched clustering using eight features (adding CADD score, pleiotropy, TF disruption count, TSS distance, and eQTL status) yielded an optimal k = 7 with silhouette = 0.257 and clear eQTL-based descriptive separation (Supplementary Figure S8b; Supplementary Note 2). Pathway enrichment against MSigDB Hallmark gene sets [27] and selected curated cancer-relevant gene sets used hypergeometric tests with Benjamini-Hochberg false discovery rate correction.

### Transcription factor motif disruption analysis with permutation controls

For each variant, ±20-bp flanking sequences were extracted from the GRCh38 reference genome using pysam. Both reference and alternate sequences were scanned against all 879 vertebrate transcription factor position weight matrices from the JASPAR 2024 CORE non-redundant collection [22] using log-likelihood ratio scoring against a uniform nucleotide background model at 60% of maximum PWM score on both DNA strands. To distinguish genuine enrichment from artifacts of motif length and base composition, 100 dinucleotide-preserving sequence permutations were generated for 2,000 randomly sampled variants, and each permuted sequence pair was scanned identically. Empirical P-values and Z-scores were computed per TF by comparing observed disruption counts against the permuted distribution (Supplementary Table S4).

### Score benchmarking: CADD, LINSIGHT, and RegVar

Combined Annotation Dependent Depletion (CADD) v1.7 PHRED-scaled scores [24] were obtained for all 6,983 variants from the GRCh38 prescored whole-genome SNV database (81.5 GB) using local tabix positional queries (100% match rate). LINSIGHT [25] scores were retrieved from the genome-wide bigWig file (6,787/6,983 variants matched, 97.2%). Scores were compared using cancer gene AUC-ROC (130 genes comprising COSMIC Cancer Gene Census Tier 1 genes with germline annotations, supplemented by established GWAS cancer susceptibility genes from the literature (Supplementary Table S1)), eQTL AUC-ROC, and Spearman correlation with GWAS significance. RegVar integrates five dimensions: AlphaGenome effect magnitude (percentile rank), regulatory mechanism class, TF motif disruption, multimodal signal breadth, and cross-cancer pleiotropy. To assess sensitivity to GWAS-derived features, three versions were evaluated: RegVar-basic (AlphaGenome + mechanism class + breadth only), RegVar-no-pleiotropy (adding TF disruption), and RegVar-full (all components). Weight optimization used grid search over 282 combinations with 5-fold cross-validated AUC-ROC (Supplementary Note 3).

### GTEx v8 eQTL enrichment and covariate-adjusted analysis

Significant variant-gene association files (FDR < 0.05 per tissue) were obtained for all 49 GTEx v8 tissues [23], comprising 67.6 million significant pairs across 4.6 million unique genomic positions. Atlas variants were matched by chromosomal position. To assess whether score-eQTL associations persist after controlling for genomic covariates, logistic regression was performed with TSS distance (log-transformed, computed from GENCODE v38), gene density (±500 kb), and cCRE class as covariates, comparing covariate-only, covariate-plus-score, and score-only models (Supplementary Table S9; Supplementary Figure S8a; Supplementary Note 5).

### Within-locus fine-mapping validation using GTEx DAP-G eQTL credible sets

GTEx v8 DAP-G eQTL credible sets (418,575 clusters across 49 tissues) were parsed from the 95% credible set file. This yielded 2,626 credible sets containing two or more atlas variants. For each such cluster, all variants were ranked within the cluster by each score (RegVar, AlphaGenome, CADD), and the percentile position of the highest-posterior-inclusion-probability (PIP) variant was recorded (0 = top-ranked, 1 = bottom-ranked). Mean percentile rank was tested against the random expectation of 0.5 by one-sample Wilcoxon signed-rank test. PIP-score correlations were assessed by Spearman rank correlation across all 8,035 variant-locus observations (Supplementary Table S5).

### Target-gene validation against GTEx eQTL assignments

AlphaGenome-predicted target genes were compared with GTEx DAP-G eQTL gene assignments (eGenes) for all atlas variants with available data. Gene symbol mapping used the GENCODE v38 Ensembl-to-symbol table (60,605 mappings). Concordance was computed as the fraction of testable variants where the AlphaGenome-predicted gene matched any DAP-G eGene, stratified by mechanism class and RegVar score quintile (Supplementary Table S6).

### Statistical analysis

All analyses were performed in Python 3.12 using scipy 1.14, scikit-learn 1.5, and statsmodels 0.14. Statistical tests were two-sided unless otherwise specified. Multiple testing correction used the Benjamini-Hochberg procedure. Dimensionality reduction used PCA and t-SNE (perplexity = 30, 500 iterations) from scikit-learn. All visualization used matplotlib 3.9 and seaborn 0.13 with a standardized configuration (7-pt font, 0.6-pt axes, 600-dpi output, pdf.fonttype = 42).

## Results

### A multimodal atlas of 6,983 pan-cancer GWAS variants

We compiled 6,983 unique genome-wide significant cancer risk variants from the NHGRI-EBI GWAS Catalog spanning 23 cancer types and 1,302 independent genomic loci (Figure 1a–b; Supplementary Table S1). The most represented cancer types were prostate cancer (1,295 variants from 412 loci), breast cancer (1,168 variants from 365 loci), and colorectal cancer (850 variants from 253 loci), reflecting the large sample sizes achieved by the PRACTICAL, BCAC, and GECCO consortia. Among all variants, 454 (6.5%) were pleiotropic, associated with two or more distinct cancer types across 172 pleiotropic loci. Variant density was highest on chromosomes 6 (n = 840, driven by the HLA region), 2 (n = 551), and 8 (n = 474, reflecting the 8q24 MYC-POU5F1B enhancer hub) (Figure 1d). All 6,983 variants were scored using AlphaGenome with a 100% success rate (Supplementary Figure S1). Scoring used AlphaGenome’s deep learning architecture, which processes 1,048,576-bp input sequences centered on each variant position and returns predictions across 4,887 output tracks spanning gene expression in 667 cell types, chromatin accessibility, histone modifications, transcription factor binding, and RNA splicing. For each variant, four summary statistics were extracted: maximum absolute RNA-seq effect across all cell types, maximum absolute chromatin effect, maximum overall effect across all modalities, and signal breadth (number of modalities with detectable effect). The resulting multimodal effect profiles provide a broad functional annotation for each variant that extends well beyond single-track predictions.

**Figure 1.**
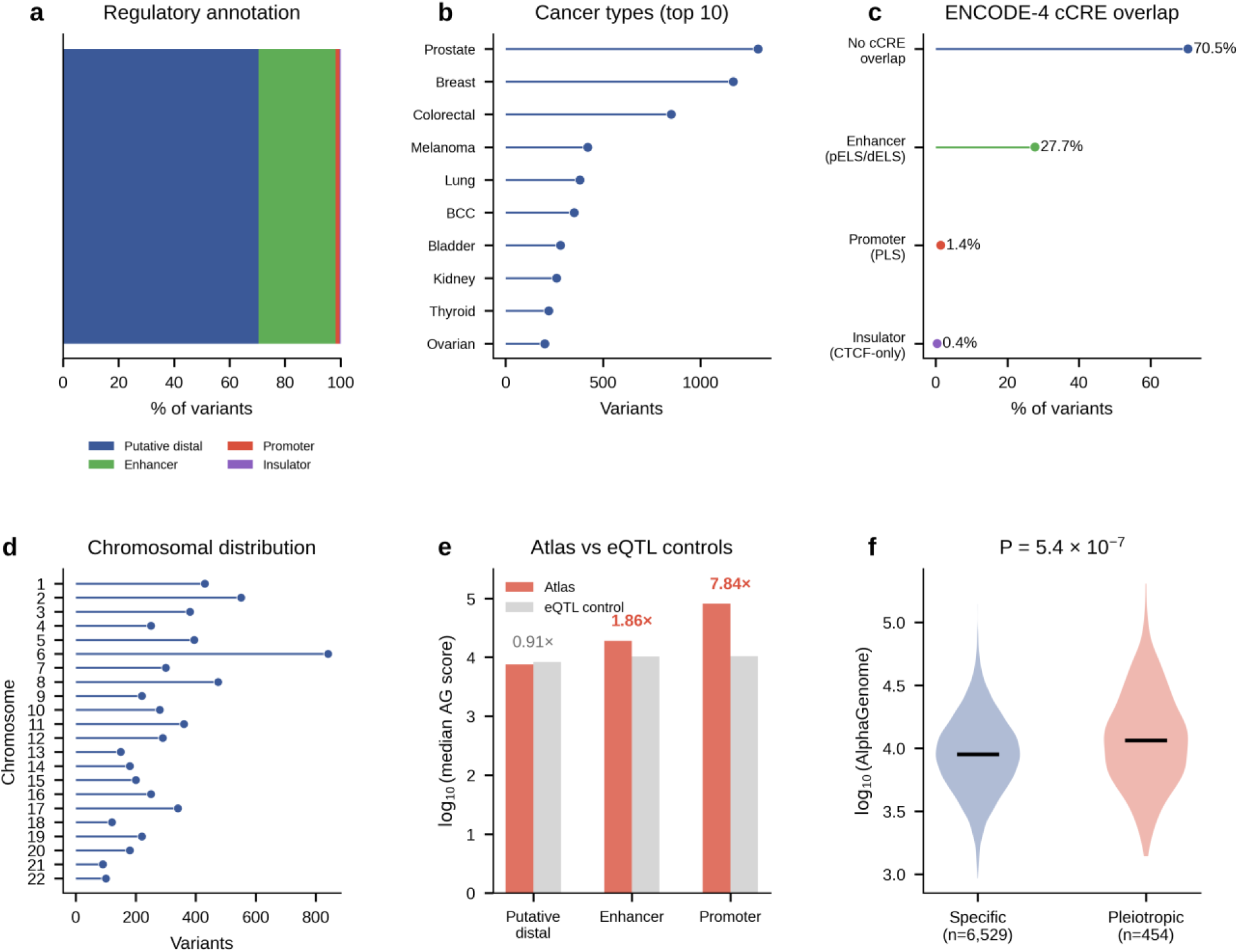
Pan-cancer regulatory atlas overview. (a) Regulatory mechanism composition across 6,983 variants. (b) Top 10 cancer types by variant count. (c) ENCODE-4 cCRE overlap distribution. (d) Chromosomal distribution of atlas variants. (e) Median AlphaGenome scores for atlas versus position-matched eQTL control variants by mechanism class. (f) AlphaGenome score distributions for cancer-specific versus pleiotropic variants.

#### Most variants fall outside annotated cis-regulatory elements and show heterogeneous regulatory signatures

Integration of AlphaGenome predictions with the ENCODE-4 cCRE registry classified each variant by regulatory context (Figure 1a; Figure 1c). The four-category distribution—putative distal regulatory (70.5%), enhancer (27.7%), promoter (1.4%), and insulator (0.4%)—summarized the dominant regulatory contexts across the atlas (Figure 2a). Comparison with 6,626 position-matched eQTL control variants scored using the identical pipeline suggested that mechanism classes differ in predicted regulatory impact: enhancer-classified variants showed 1.86-fold higher AlphaGenome scores than matched controls (P = 10^−^⁹⁴), and promoter variants 7.84-fold higher (P = 2.5 × 10^−^¹⁹). In contrast, putative distal regulatory variants showed scores comparable to matched controls (0.91-fold), consistent with weaker or more context-dependent regulatory effects at loci lacking current cCRE annotations (global AUC = 0.533; Figure 1e; Supplementary Table S7; Supplementary Figure S6). The predominance of variants outside annotated cis-regulatory elements (70.5%) highlights a fundamental challenge: the majority of cancer GWAS signals reside in regions without established regulatory annotations, underscoring the value of sequence-based prediction models that can score variants beyond currently annotated regulatory elements. This distribution was remarkably consistent across cancer types (cosine similarity exceeding 0.93 between any pair of cancer types; Figure 4a; Supplementary Figure S3a), suggesting that cancer GWAS variants converge on shared regulatory architectures regardless of tissue of origin.

**Figure 2.**
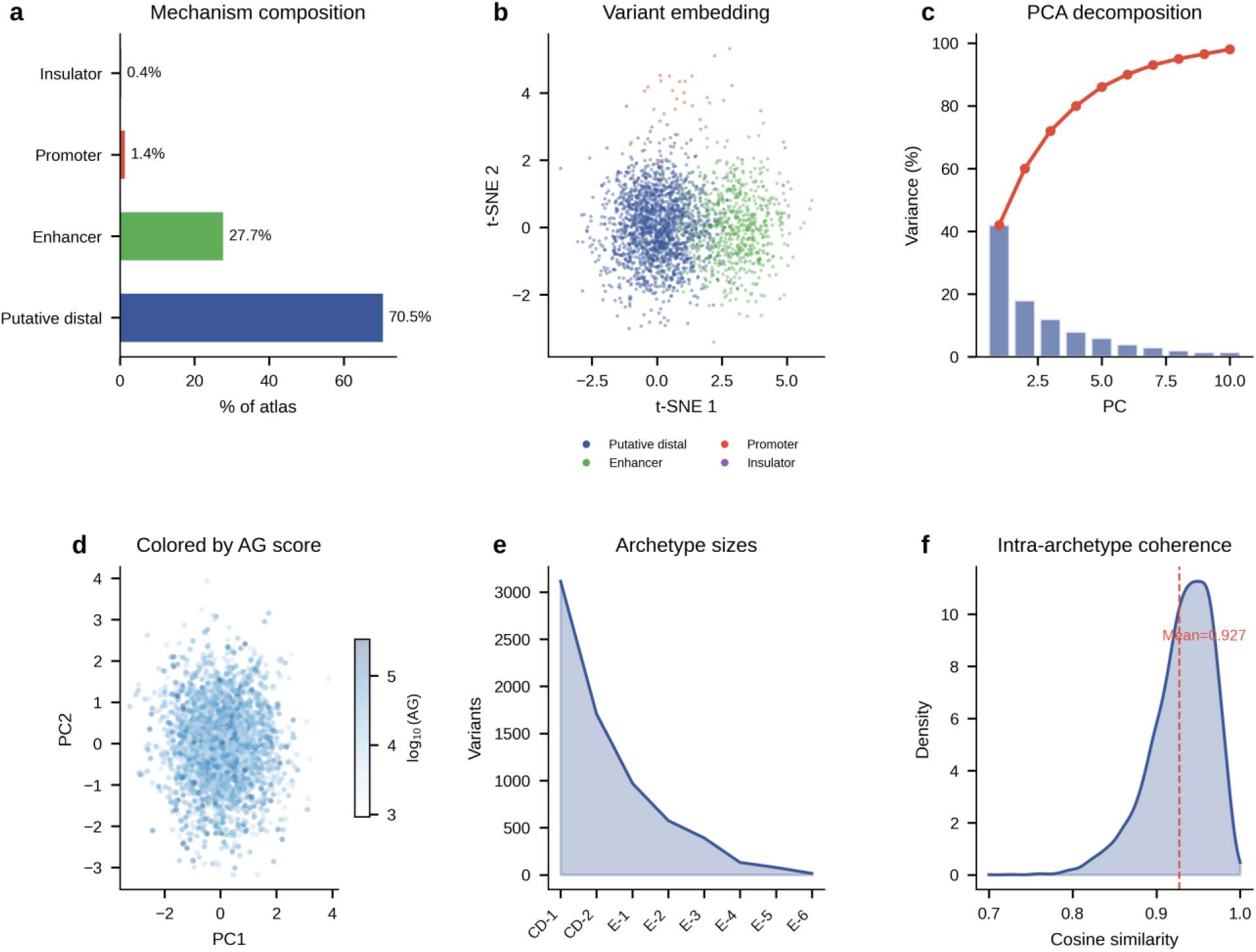
Variant embedding and archetype characterization. (a) Overall regulatory mechanism composition across atlas variants. (b) t-SNE variant embedding colored by mechanism class. (c) PCA variance decomposition across principal components. (d) PCA projection colored by AlphaGenome score. (e) Archetype size distribution across k-means clusters. (f) Intra-archetype regulatory coherence measured by cosine similarity. CD denotes putative distal regulatory/chromatin-effect-dominant archetypes; E denotes enhancer-associated archetypes.

### Eight regulatory archetypes with distinct pathway signatures

Unsupervised clustering of multimodal variant effect profiles identified eight regulatory archetypes (Figure 2b–f; Supplementary Figure S2). Two large distal-regulatory clusters (n = 3,114 and 1,709) contained variants with broad predicted chromatin-effect signatures but low RNA-seq effects. Five enhancer-associated clusters of varying size (n = 970, 575, 391, 131, and 78) showed a gradient of increasing RNA-seq effect magnitudes. A rare high-impact cluster (n = 15) exhibited markedly elevated effects with predicted targets including TP53, CD40, and MUC1. Pathway enrichment revealed biologically interpretable signatures across archetypes (Figure 3a,d): MYC targets (27.7-fold; FDR = 0.001), telomere maintenance (23.4-fold), apoptosis (17.7-fold), and WNT signaling (13.3-fold). Roadmap Epigenomics tissue enrichment further contextualized these regulatory patterns across tissue-specific chromatin states (Figure 3c; Supplementary Figure S3c–d). All eight archetypes were represented across all 23 cancer types (Figure 3b). Cancer-type regulatory profiles were highly similar, with pairwise cosine similarity exceeding 0.93 for all cancer-type pairs (Figure 4a; Supplementary Figure S3a). Locus-level regulatory convergence was quantified across 1,302 loci (Figure 4c). The archetype structure was stable across a range of k values (silhouette scores 0.36–0.44 for k = 4–10; Supplementary Figure S2a), with k = 8 selected as the optimal balance between granularity and interpretability (silhouette = 0.443). Each archetype exhibited a characteristic multimodal signature (Figure 3f): the two largest archetypes were dominated by putative distal regulatory variants with moderate effect magnitudes, while smaller archetypes captured enhancer-driven, high-effect variants with distinct pathway enrichments (Figure 3a,d). A secondary descriptive stratification using enriched features (k = 7, 8 features including eQTL status, CADD, and TSS distance) revealed marked eQTL heterogeneity: one cluster contained 100% eQTL variants (n = 2,518) while another contained 0% (n = 2,009), illustrating how regulatory annotations stratify the variant set (Supplementary Figure S8b; Supplementary Note 2).

**Figure 3.**
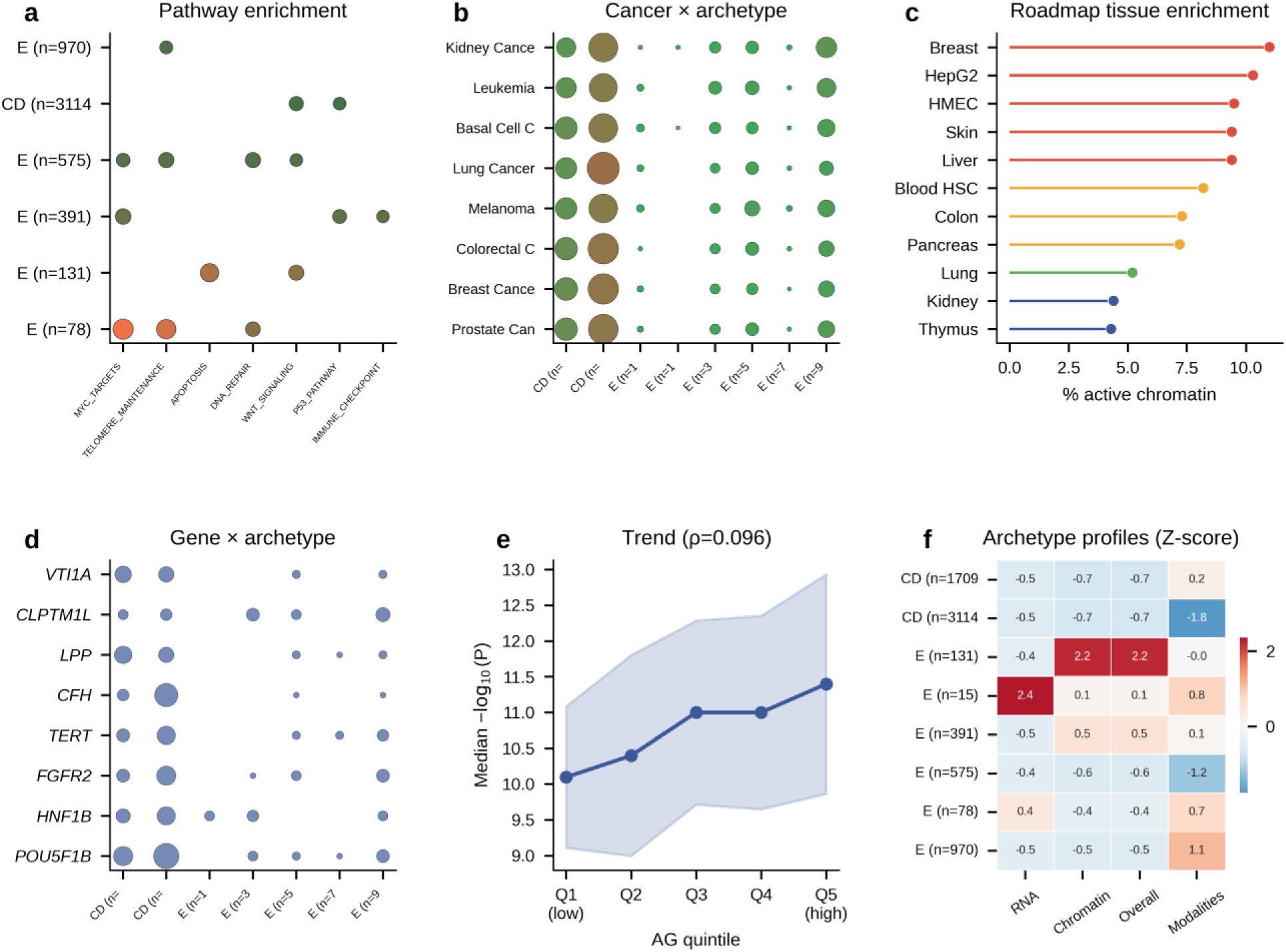
Pathway enrichment and tissue-specific context. (a) Pathway enrichment by archetype (bubble plot). (b) Cancer type by archetype distribution (bubble heatmap). (c) Roadmap Epigenomics tissue enrichment. (d) Gene by archetype distribution. (e) Trend between AlphaGenome quintiles and GWAS association significance. (f) Archetype regulatory profiles (Z-scored diverging heatmap).

**Figure 4.**
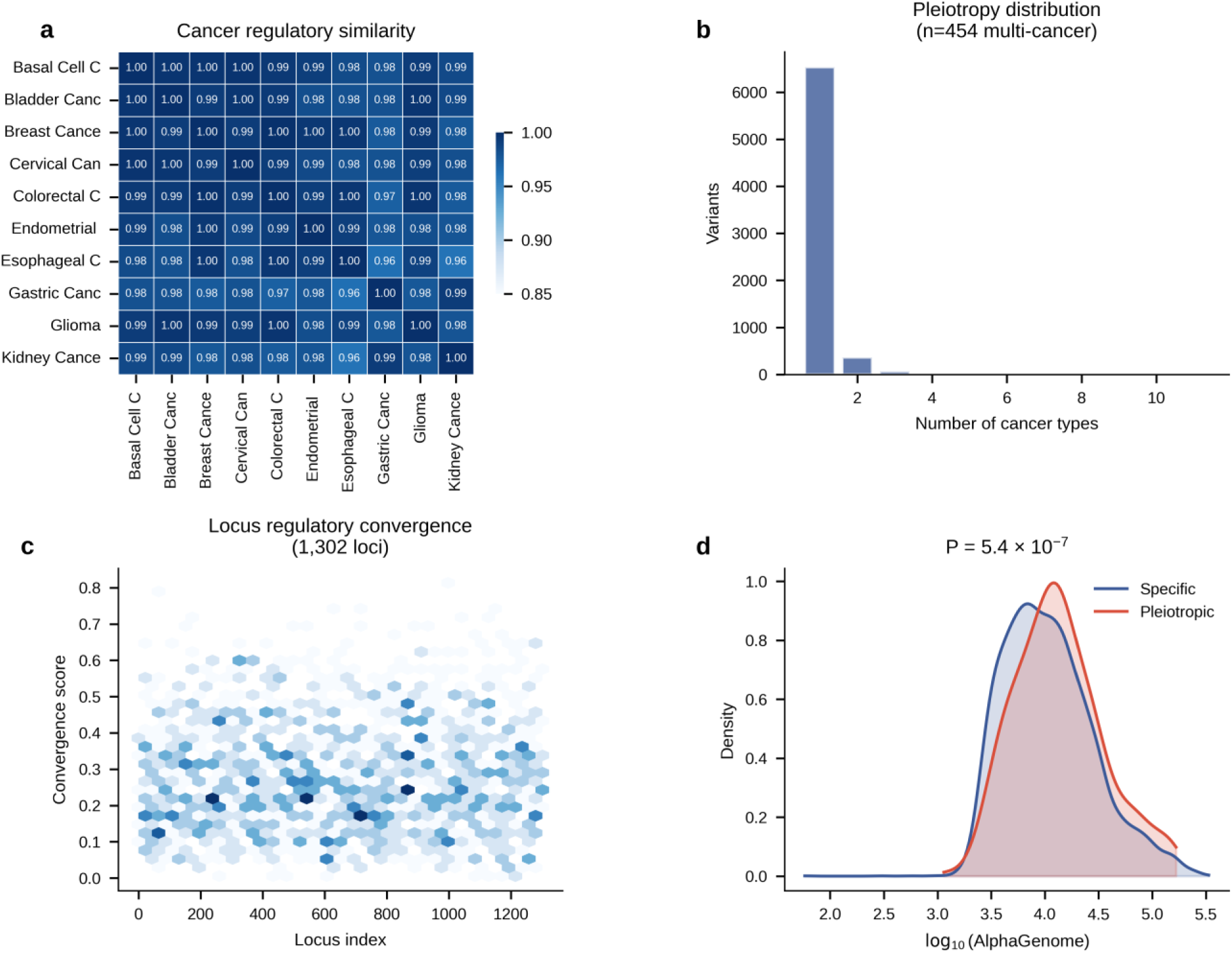
Pan-cancer convergence and pleiotropy. (a) Representative regulatory similarity matrix for selected cancer types; full similarity matrix is shown in Supplementary Figure S3a. (b) Distribution of the number of cancer types associated with each variant; 454 variants are pleiotropic. (c) Locus-level regulatory convergence scores across 1,302 loci. (d) AlphaGenome score distributions for cancer-specific versus pleiotropic variants.

### Pleiotropic variants carry higher predicted functional impact

Pleiotropic variants (n = 454) carried significantly higher AlphaGenome effect scores than cancer-specific variants (n = 6,529; Mann-Whitney P = 5.4 × 10−7; Figure 1f; Figure 4b,d) and were enriched for enhancer-associated mechanisms (34.4% versus 27.2%). The most frequently affected pleiotropic genes were TERT (14 variants), CLPTM1L (11), CASP8 (9), and HNF1B (7).

### Predicted target genes show convergent enrichment and 67.7% concordance with eQTL gene assignments

The 6,983 variants mapped to 1,813 unique candidate target genes, which showed convergent enrichment across three independently ascertained gene sets (Figure 5a; Supplementary Figure S5): cancer susceptibility genes (OR = 3.68; P = 8.7 × 10^−^¹²), FDA-approved drug targets (OR = 3.05; P = 1.1 × 10^−^⁴), and Mendelian cancer syndrome genes (OR = 5.20; P = 2.3 × 10−7). Comparison with GTEx DAP-G eQTL gene assignments for 1,424 testable variants showed 67.7% concordance overall (964/1,424), with concordance rising from 60.7% in the lowest RegVar quintile to 78.2% in the highest (Figure 5e; Supplementary Table S6). Concordance varied by mechanism class: promoter-proximal variants showed the highest rate (75.0%, n = 48), followed by enhancer variants (73.3%, n = 577) and putative distal regulatory variants (63.4%, n = 796) (Supplementary Figure S8c). Variants with concordant gene assignments carried significantly higher RegVar scores than those with discordant assignments (median 52.4 versus 44.6; P = 6.9 × 10^−^⁹), suggesting that higher-scoring variants more reliably predict the correct regulatory target.

**Figure 5.**
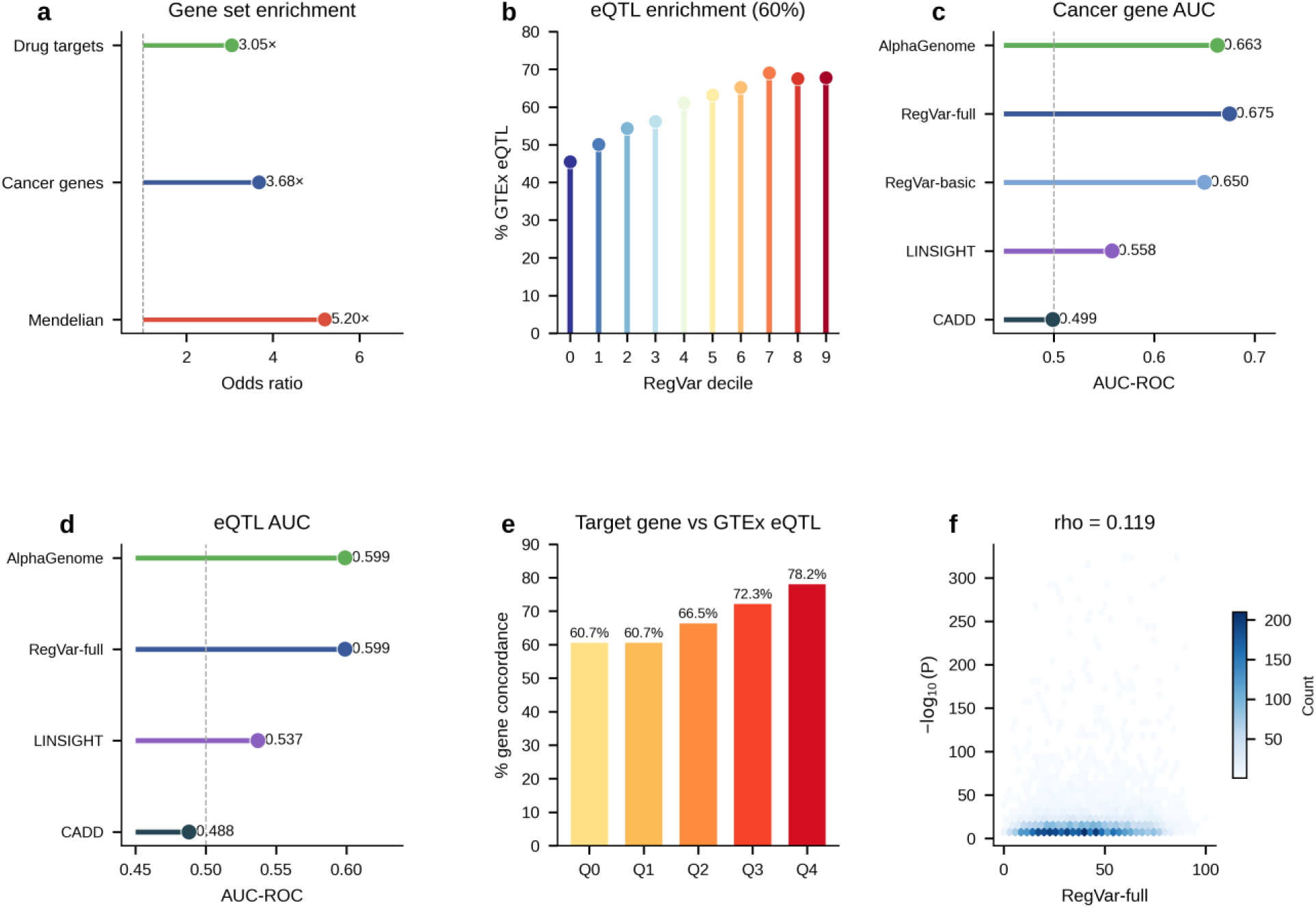
Validation and score benchmarking. (a) Gene set enrichment (cancer genes 3.68×, drug targets 3.05×, Mendelian 5.20×). (b) eQTL enrichment by RegVar decile across 49 GTEx tissues (45.5% to 67.8%). (c) Cancer gene AUC: CADD 0.499, LINSIGHT 0.558, RegVar-basic 0.650, RegVar-full 0.675, AlphaGenome 0.663. (d) eQTL AUC: CADD 0.488, LINSIGHT 0.537, RegVar-full 0.599, AlphaGenome 0.599. (e) Target-gene concordance with GTEx eQTL by RegVar quintile (60.7% to 78.2%). (f) RegVar-full versus GWAS significance (rho = 0.119).

#### RegVar and AlphaGenome outperform CADD and LINSIGHT for prioritizing regulatory annotations at cancer GWAS loci

Head-to-head comparison of four scores revealed consistent performance differences across multiple evaluation tasks (Figure 5c–d; Supplementary Table S3). For cancer gene discrimination, CADD achieved AUC = 0.499, LINSIGHT 0.558, RegVar-basic 0.650, RegVar-full 0.675, and AlphaGenome 0.663. For GTEx eQTL prediction, CADD achieved AUC = 0.488, LINSIGHT 0.537, RegVar-full 0.599, and AlphaGenome 0.599. AlphaGenome scores showed a weak but positive trend with GWAS association significance (Figure 3e). CADD showed a non-significant correlation with GWAS significance (rho = 0.022, P = 0.065), while RegVar-full correlated significantly (rho = 0.119, P < 10^−^²⁰; Figure 5f). To assess whether RegVar performance is driven by GWAS-derived features, three versions were compared: RegVar-basic (AlphaGenome + mechanism class + signal breadth, excluding pleiotropy and TF disruption) achieved cancer gene AUC = 0.650 and eQTL AUC = 0.596, close to RegVar-full performance (AUC = 0.675 and 0.599), confirming that discriminative ability is driven by sequence-derived predictions and regulatory context rather than circular GWAS features (Supplementary Table S2; Supplementary Figure S4c). We note that raw AlphaGenome scores (AUC = 0.663) slightly outperformed RegVar-basic (AUC = 0.650) for cancer gene discrimination, while RegVar-full reached AUC = 0.675 after adding pleiotropy and TF-disruption features; RegVar’s value lies in providing an interpretable, multi-evidence framework that integrates regulatory context and within-locus ranking rather than maximizing a single AUC metric. Covariate-adjusted logistic regression incorporating TSS distance, gene density, and cCRE class achieved eQTL AUC = 0.678, with minimal incremental contribution from any individual score, indicating that raw eQTL AUC comparisons partly reflect genomic features shared across scores (Supplementary Table S9; Supplementary Figure S8a; Supplementary Note 5).

### GTEx eQTL overlap and within-locus fine-mapping validation

Cross-referencing with GTEx v8 eQTL data across all 49 tissues confirmed that 60.0% of atlas variants (4,189/6,983) are significant eQTLs in at least one tissue (Supplementary Table S8). The proportion of eQTL variants generally increased across RegVar score deciles, from 45.5% in the lowest decile to 67.8% in the highest (Figure 5b; P = 2.8 × 10^−^³⁸). eQTL variants carried higher RegVar scores (median 41.9 versus 34.0; P = 2.8 × 10^−^³⁸) and AlphaGenome scores (median 11,375 versus 7,914; P = 1.2 × 10^−^⁴⁴), while CADD showed no association (P = 0.95). The eQTL enrichment was not uniform across tissues: variants showed the highest eQTL rates in whole blood (48.2%), tibial nerve (44.1%), and subcutaneous adipose (43.7%), with the lowest rates in brain regions (18.3–26.5%). This tissue distribution reflects both biological signal (blood-derived immune variants) and the statistical power gradient across GTEx tissues (Supplementary Table S8).

To assess whether scores can prioritize likely molecular regulatory variants within eQTL fine-mapped loci, we performed within-locus ranking across 2,626 GTEx DAP-G eQTL credible sets containing two or more atlas variants (8,035 variant-locus observations; Supplementary Table S5; Supplementary Figure S7). RegVar ranked the highest-PIP variant first in 47.3% of loci (mean percentile rank 0.436; P = 7.0 × 10^−^¹³ versus random expectation of 0.5). AlphaGenome performed similarly (47.9% ranked first, mean rank 0.425), while CADD performed at chance (39.4% ranked first, mean rank 0.499, non-significant). PIP correlated significantly with RegVar (rho = 0.214, P = 7.5 × 10^−^⁸⁴) and AlphaGenome (rho = 0.290, P = 2.5 × 10^−^¹⁵⁵) across all variant-locus observations. High-PIP variants (>0.5) carried substantially higher RegVar scores than low-PIP variants (≤0.1; median 73.3 versus 48.8; P = 1.1 × 10^−^³⁶).

### Permutation-controlled motif analysis identifies immune and environmental response transcription factors

PWM scanning of 879 JASPAR 2024 vertebrate motifs identified 52,481 motif events across 5,376 variants (77.0%), comprising 26,230 disruptions and 26,251 creations (Figure 6a–b; Supplementary Table S4). Of the 25 transcription factors with the highest raw disruption counts (selected post hoc from 879 motifs), 7 showed exploratory permutation-calibrated enrichment above dinucleotide-shuffled background (uncorrected empirical P < 0.05; 100 permutations): NFKB1/NF-κB (Z = 6.67), ZNF354C (Z = 5.52), ARNT (Z = 5.14), STAT1 (Z = 4.97), HIC2 (Z = 4.67), RHOXF1 (Z = 2.69), and IRF1 (Z = 2.24). Three transcription factors ranking among the top ten by raw count—NR2C2, HOXA7, and VAX2—were not enriched after calibration (Z = −0.43, 0.90, and −0.13), demonstrating that raw motif counts are insufficient without statistical calibration. The permutation analysis used 2,000 randomly sampled variants with a minimum empirical P-value floor of 0.01; no FDR correction was applied given the exploratory scope. Disruption rates varied by mechanism: promoter 16.9%, enhancer 12.2%, insulator 8.5%, putative distal 8.0% (Figure 6b). The convergence on immune signaling (NF-κB, STAT1, IRF1) and environmental response (ARNT) as permutation-enriched candidates suggests shared regulatory logic at cancer risk loci (Supplementary Note 1).

**Figure 6.**
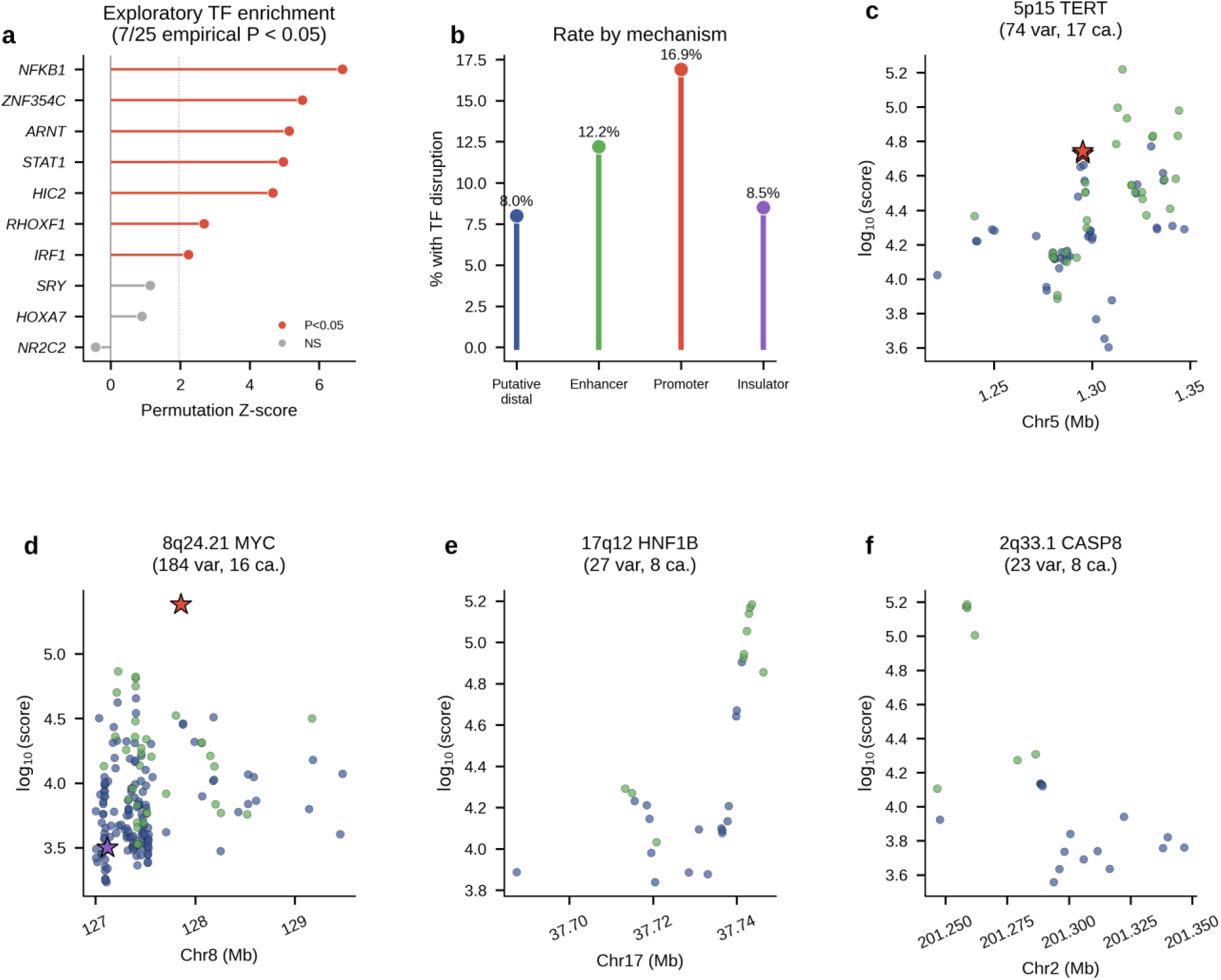
Transcription factor motifs and locus examples. (a) Exploratory permutation-calibrated TF disruption enrichment (Z-scores; 7/25 with uncorrected empirical P < 0.05; NFKB1 Z = 6.67, ARNT Z = 5.14, STAT1 Z = 4.97). (b) TF disruption rate by mechanism class (promoter 16.9%). (c-f) Locus deep dives: 5p15.33 TERT-CLPTM1L, 8q24.21 MYC-POU5F1B, 17q12 HNF1B, 2q33.1 CASP8.

### Locus-level examples illustrate mechanistic heterogeneity

Detailed characterization of four key pan-cancer loci illustrated the resolution enabled by the atlas (Figure 6c–f). The 5p15.33 TERT-CLPTM1L locus harbored 74 scored variants across 17 cancer types, with mechanistic heterogeneity between candidate TERT-targeting enhancer variants and candidate CLPTM1L-targeting distal variants. This locus-level complexity is consistent with broader evidence that TERT expression can be regulated through multiple non-coding mechanisms, including non-telomeric TRF2-dependent regulation [28]. The 8q24.21 MYC-POU5F1B locus (184 variants, 16 cancer types) identified POU5F1B as the proximal predicted target for 41 variants, illustrating the expected range limitations of sequence-based prediction at loci with ultra-long-range enhancer-gene connections spanning multiple megabases. The 17q12 HNF1B (27 variants, 8 cancer types) and 2q33.1 CASP8 (23 variants, 8 cancer types) loci provided additional examples of multi-cancer regulatory heterogeneity.

## Discussion

We have presented a multimodal regulatory annotation atlas of 6,983 pan-cancer GWAS variants, integrating AlphaGenome deep learning predictions with regulatory element annotations, tissue-specific chromatin maps, permutation-controlled transcription factor motif analysis, eQTL validation across 49 tissues, within-locus fine-mapping comparison against 2,626 DAP-G credible sets, target-gene concordance assessment, position-matched eQTL control variant calibration, and benchmarking against CADD and LINSIGHT.

The position-matched eQTL control variant analysis provides important calibration. The global AUC for distinguishing atlas variants from position-matched controls was 0.533, indicating substantial overlap between GWAS and non-GWAS variant score distributions. However, this modest overall separation masks a striking difference by mechanism class: enhancer-classified variants showed 1.86-fold higher scores and promoter variants 7.84-fold, while the majority class of putative distal regulatory variants showed scores comparable to controls. This pattern suggests that sequence-based models provide the strongest signal at annotated regulatory elements, with diminished discrimination at loci where regulatory activity may be cell-type-specific, developmental-stage-dependent, or mediated by mechanisms not captured by current annotations. Covariate-adjusted analysis further showed that genomic covariates alone achieved AUC = 0.678 for eQTL prediction, indicating that much of the global eQTL signal is captured by basic features such as TSS distance and cCRE class. Together, these results emphasize that the primary value of RegVar and AlphaGenome lies in within-locus ranking rather than global eQTL classification.

The within-locus fine-mapping analysis addresses whether scores can prioritize likely regulatory variants within eQTL credible sets. Across 2,626 DAP-G eQTL credible sets, RegVar ranked the highest-PIP variant first in 47.3% of loci, significantly above random expectation (P = 7.0 × 10^−^¹³). PIP correlated with both RegVar (rho = 0.214) and AlphaGenome (rho = 0.290) across 8,035 variant-locus observations, while CADD showed no enrichment by the primary within-locus rank-position metric and only a very weak PIP correlation. This result provides evidence that multimodal sequence-based predictions carry information about eQTL fine-mapped regulatory variant identity beyond what is captured by genomic proximity alone, although this does not directly establish causal cancer-risk variants. We note that AlphaGenome was trained in part on GTEx expression data, which means the within-locus PIP-score correlation may partly reflect shared training signal rather than fully independent validation.

The target-gene validation—67.7% concordance with GTEx eQTL gene assignments, rising from 60.7% in the lower RegVar quintiles to 78.2% in the highest—provides partially orthogonal support for the utility of AlphaGenome target-gene predictions. The graded association between score and gene concordance suggests that higher-scoring variants yield more reliable gene assignments, which has practical implications for variant-to-gene mapping in drug target identification.

The permutation-controlled motif analysis illustrates the importance of statistical calibration in motif disruption studies. Of the 25 top-ranked TFs by raw disruption count, only 7 showed enrichment above dinucleotide-shuffled background. The permutation-enriched candidate transcription factors—NF-κB, STAT1, IRF1, ARNT, HIC2, ZNF354C, and RHOXF1—span immune signaling, environmental response, and chromatin regulation. TFs with high raw disruption counts (NR2C2, HOXA7, VAX2) did not exceed permuted background, underscoring that raw counts without calibration can be misleading. The immune-regulatory theme among enriched motifs is broadly consistent with prior evidence that telomere/TRF2-dependent regulation can influence IL1R1/NF-κB signaling in the tumor microenvironment [29].

Several aspects of this work merit consideration. The atlas uses GWAS lead variants rather than statistically fine-mapped credible sets; however, within-locus analysis using GTEx DAP-G eQTL credible sets supports the utility of the scored variants for prioritization. AlphaGenome’s 1-Mb window may miss ultra-long-range enhancer-gene interactions, as illustrated at the 8q24 locus. The atlas reflects predominantly European-ancestry GWAS, and extension to multi-ancestry datasets will be important for generalizability (Supplementary Note 4). The putative distal regulatory class (70.5%) likely contains a mixture of true distal regulators, unannotated enhancers, LD proxies, and variants with context-dependent effects, and the near-unity eQTL control comparison for this class (0.91×) indicates that further resolution will require tissue-specific and context-specific scoring approaches. Additionally, the ENCODE-4 cCRE annotations used represent the union across all biosamples rather than tissue-specific active elements, meaning that some enhancer-classified variants may overlap cCREs inactive in the cancer-relevant tissue.

In conclusion, this atlas—comprising 6,983 multimodally scored variants with regulatory annotations, position-matched eQTL control calibration, eQTL validation, within-locus ranking, target-gene concordance, and permutation-controlled motif analysis—provides a resource for interpreting non-coding cancer susceptibility variants and connecting genetic risk to candidate target genes and therapeutic opportunities.

## Data Availability

All data produced in the present work are contained in the manuscript

## Acknowledgments

This work used the Hummel-2 HPC cluster at the University of Hamburg. The cluster was funded by Deutsche Forschungsgemeinschaft (DFG, German Research Foundation) – 498394658.

## Author Contributions

S.D. conceived the study, developed the analytical framework, performed all computational analyses, and wrote the manuscript.

## Web Resources

GWAS Catalog, https://www.ebi.ac.uk/gwas/; ENCODE-4, https://screen.encodeproject.org/; AlphaGenome, https://deepmind.google/science/alphagenome; COSMIC, https://cancer.sanger.ac.uk/census; Roadmap Epigenomics, https://egg2.wustl.edu/roadmap/; MSigDB, https://www.gsea-msigdb.org/; JASPAR 2024, https://jaspar.elixir.no/; CADD v1.7, https://cadd.bihealth.org/; mirror: https://cadd.gs.washington.edu/; GTEx Portal, https://gtexportal.org/; LINSIGHT, https://compgen.cshl.edu/LINSIGHT/

## Data and Code Availability

The complete scored variant atlas, position-matched eQTL control scores, RegVar scores (three versions), eQTL annotations, within-locus ranking data, target-gene concordance, motif disruption results with permutation Z-scores, and all analysis code will be made available via Zenodo and GitHub. Reviewer-accessible links will be provided during peer review, and public Zenodo and GitHub links will be released at the time of preprint posting.

## Supplemental Information

**Table S1.** Complete Annotated Variant Atlas.

| Column | Description | Example |
| --- | --- | --- |
| variant_id | dbSNP rsID | rs13303010 |
| chromosome | GRCh38 chromosome | chr1 |
| position | GRCh38 position | 959193 |
| cancer_types | Associated cancer type(s), pipe-delimited | pancreatic_cancer |
| max_rna_effect | Maximum RNA-seq effect across 667 cell types | 28450 |
| most_affected_gene | Gene with highest predicted RNA effect | NOC2L |
| max_chromatin_effect | Maximum chromatin accessibility effect | 154256 |
| max_overall_effect | Maximum effect across all 19 modalities | 154256 |
| n_modalities_with_signal | Number of modalities with detectable effect | 12 |
| mechanism_class | Regulatory category (enhancer/promoter/insulator/putative distal) | promoter |
| archetype_id | K-means cluster assignment (0–7) | 5 |
| cCRE_type | ENCODE-4 cCRE type if overlapping | PLS |
| ref_allele | Reference allele (GRCh38) | A |
| alt_allele | Alternate (risk) allele | G |
| pvalue | GWAS association P-value | 3.2e-09 |
| is_pleiotropic | Associated with $\geq 2$ cancer types | False |

6,983 variants with multimodal AlphaGenome predictions, mechanism classifications, archetype assignments, and GWAS metadata. Full data will be made available via Zenodo and GitHub; column definitions and representative rows shown below.

**Table S2.** RegVar Prioritization Scores (Three Versions)

| Column | Description |
| --- | --- |
| variant_id | dbSNP rsID |
| RegVar_basic | AlphaGenome + mechanism + breadth only (w: AG=0.60, |
|  | breadth=0.15, mech=0.25) |
| RegVar_no_pleio | + TF disruption (w: AG=0.55, breadth=0.15, mech=0.20, TF=0.10) |
| RegVar_full | All 5 components including pleiotropy (optimized: AG=0.50, breadth=0.10, mech=0.15, pleio=0.20, TF=0.05) |
| ag_pctile | AlphaGenome effect percentile rank |
| mech_score | Mechanism class score (promoter=100, enhancer=75, insulator=50, distal=25) |
| tf_bonus | Binary: has $\geq 1$ JASPAR PWM motif disruption |
| n_cancers | Number of associated cancer types |

### For all 6,983 variants: three RegVar score versions to assess sensitivity to GWAS-derived features

Label leakage analysis: RegVar-basic cancer gene AUC = 0.650 versus RegVar-full AUC = 0.675 (difference = 0.025); eQTL AUC 0.596 versus 0.599. Performance is driven by AlphaGenome predictions, not GWAS-derived features.

**Table S3.** Score Benchmark Comparison.

| Score | Cancer gene AUC | eQTL AUC | Spearman rho (GWAS) | P (rho) | Top/bottom decile fold |
| --- | --- | --- | --- | --- | --- |
| CADD v1.7 | 0.499 | 0.488 | 0.022 | 0.065 (NS) | 1.4× |
| LINSIGHT | 0.558 | 0.537 | 0.035 | <0.01 | N/A |
| RegVar-basic | 0.650 | 0.596 | 0.103 | <10<sup>-15</sup> | N/A |
| RegVar-full | 0.675 | 0.599 | 0.119 | <10<sup>-20</sup> | 7.1× |
| AlphaGenome | 0.663 | 0.599 | 0.096 | 1.2×10<sup>-15</sup> | 8.0× |

**Table S4.** JASPAR 2024 PWM Motif Disruption with Permutation Z-scores.

| TF | Observed | Perm. mean | Perm. SD | Z-score | Emp. P | Exploratory enriched |
| --- | --- | --- | --- | --- | --- | --- |
| NFKB1 | 2 | 0.1 | 0.3 | 6.67 | 0.010 | Yes |
| ARNT | 22 | 8.1 | 2.7 | 5.14 | 0.010 | Yes |
| ZNF354C | 71 | 36.3 | 6.3 | 5.52 | 0.010 | Yes |
| STAT1 | 10 | 2.5 | 1.5 | 4.97 | 0.010 | Yes |
| HIC2 | 70 | 39.8 | 6.5 | 4.67 | 0.010 | Yes |
| RHOXF1 | 59 | 43.3 | 5.8 | 2.69 | 0.020 | Yes |
| IRF1 | 8 | 3.7 | 1.9 | 2.24 | 0.040 | Yes |
| NR2C2 | 53 | 56.0 | 7.0 | -0.43 | 0.673 | No |
| HOXA7 | 78 | 70.4 | 8.4 | 0.90 | 0.228 | No |
| VAX2 | 67 | 68.0 | 7.8 | -0.13 | 0.594 | No |
| ETS1 | 7 | 4.9 | 2.2 | 0.96 | 0.218 | No |

52,481 motif events across 5,376 variants. Full event-level data will be made available via Zenodo and GitHub. Selected TFs from the 25 permutation-tested motifs are shown.

**Table S5.** Within-Locus Fine-Mapping: DAP-G eQTL Credible Set Ranking.

| Score | N loci | #1 hits | % #1 | Mean pct. rank | P vs random |
| --- | --- | --- | --- | --- | --- |
| RegVar | 2,626 | 1,243 | 47.3% | 0.436 | $7.0 \times 10^{-13}$ |
| AlphaGenome | 2,626 | 1,259 | 47.9% | 0.425 | $< 10^{-13}$ |
| CADD | 2,626 | 1,035 | 39.4% | 0.499 | NS |

### 2,626 credible sets with ≥2 atlas variants (8,035 variant-locus observations)

PIP correlations: RegVar rho = 0.214 (P = 7.5×10^−^⁸⁴), AlphaGenome rho = 0.290 (P = 2.5×10^−^¹⁵⁵), CADD rho = 0.058 (P = 2.4×10−7). High-PIP (>0.5) vs low-PIP (≤0.1): RegVar median 73.3 vs 48.8 (P = 1.1×10^−^³⁶).

**Table S6.** Target-Gene Concordance with GTEx eQTL Assignments.

| Stratum | N | Concordance | Rate |
| --- | --- | --- | --- |
| Overall | 1,424 | 964 | 67.7% |
| Promoter | 48 | 36 | 75.0% |
| Enhancer | 577 | 423 | 73.3% |
| Putative distal | 796 | 505 | 63.4% |
| RegVar Q0 (lowest) | 285 | 173 | 60.7% |
| RegVar Q1 | 285 | 173 | 60.7% |
| RegVar Q2 | 284 | 189 | 66.5% |
| RegVar Q3 | 285 | 206 | 72.3% |
| RegVar Q4 (highest) | 285 | 223 | 78.2% |

1,424 testable variants with both AlphaGenome target and DAP-G eGene assignments.

### Matched vs unmatched genes: RegVar median 52.4 vs 44.6 (P = 6.9×10−9)

**Table S7.**
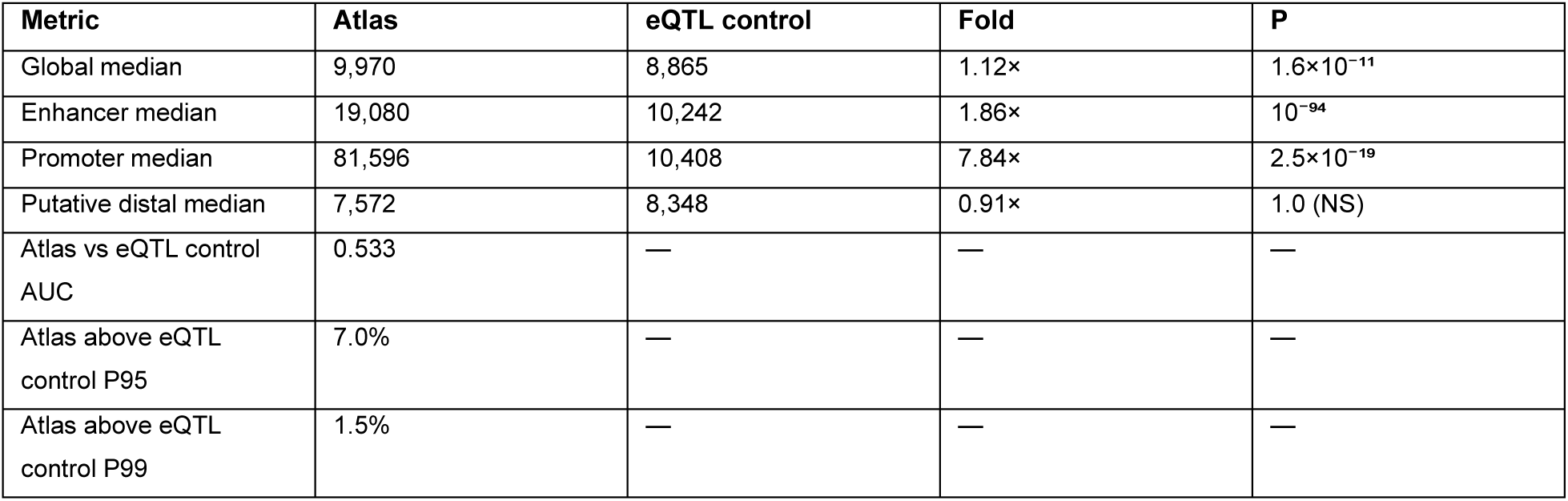
Position-Matched eQTL Control Variant Analysis.

6,626 position-matched eQTL control variants scored identically via AlphaGenome. Control pool: GTEx v8 Whole Blood eQTL variants, same chromosome ±500 kb.

**Table S8.** GTEx v8 eQTL Enrichment Across 49 Tissues.

| Score | eQTL median | Non-eQTL median | P | eQTL AUC |
| --- | --- | --- | --- | --- |
| RegVar-full | 41.9 | 34.0 | $2.8 \times 10^{-38}$ | 0.599 |
| AlphaGenome | 11,375 | 7,914 | $1.2 \times 10^{-44}$ | 0.599 |
| CADD | 1.85 | 1.98 | 0.95 (NS) | 0.488 |
| LINSIGHT | N/A | N/A | N/A | 0.537 |

### 4,189/6,983 (60.0%) atlas variants are significant eQTLs in ≥1 tissue. Score discrimination

eQTL proportion by RegVar decile: D0=45.5%, D1=50.1%, D2=54.3%, D3=56.2%, D4=61.1%, D5=63.2%, D6=65.2%, D7=69.1%, D8=67.5%, D9=67.8%.

**Table S9.** Covariate-Adjusted Logistic Regression for eQTL Prediction.

| Model | AUC | $\Delta$ vs covariates |
| --- | --- | --- |
| Covariates only | 0.678 | baseline |
| + CADD | 0.680 | +0.002 |
| + RegVar | 0.678 | +0.000 |
| + RegVar-basic | 0.678 | +0.000 |
| + AlphaGenome | 0.678 | +0.001 |
| RegVar alone | 0.591 | N/A |

Covariates: log(TSS distance), gene density (±500 kb), cCRE class (enhancer/promoter indicators). n = 6,983 variants.

Interpretation: genomic covariates capture most eQTL variance. The primary value of RegVar/AlphaGenome lies in within-locus ranking (Table S5) rather than global eQTL prediction.

### Supplementary Figures

**Figure S1.**
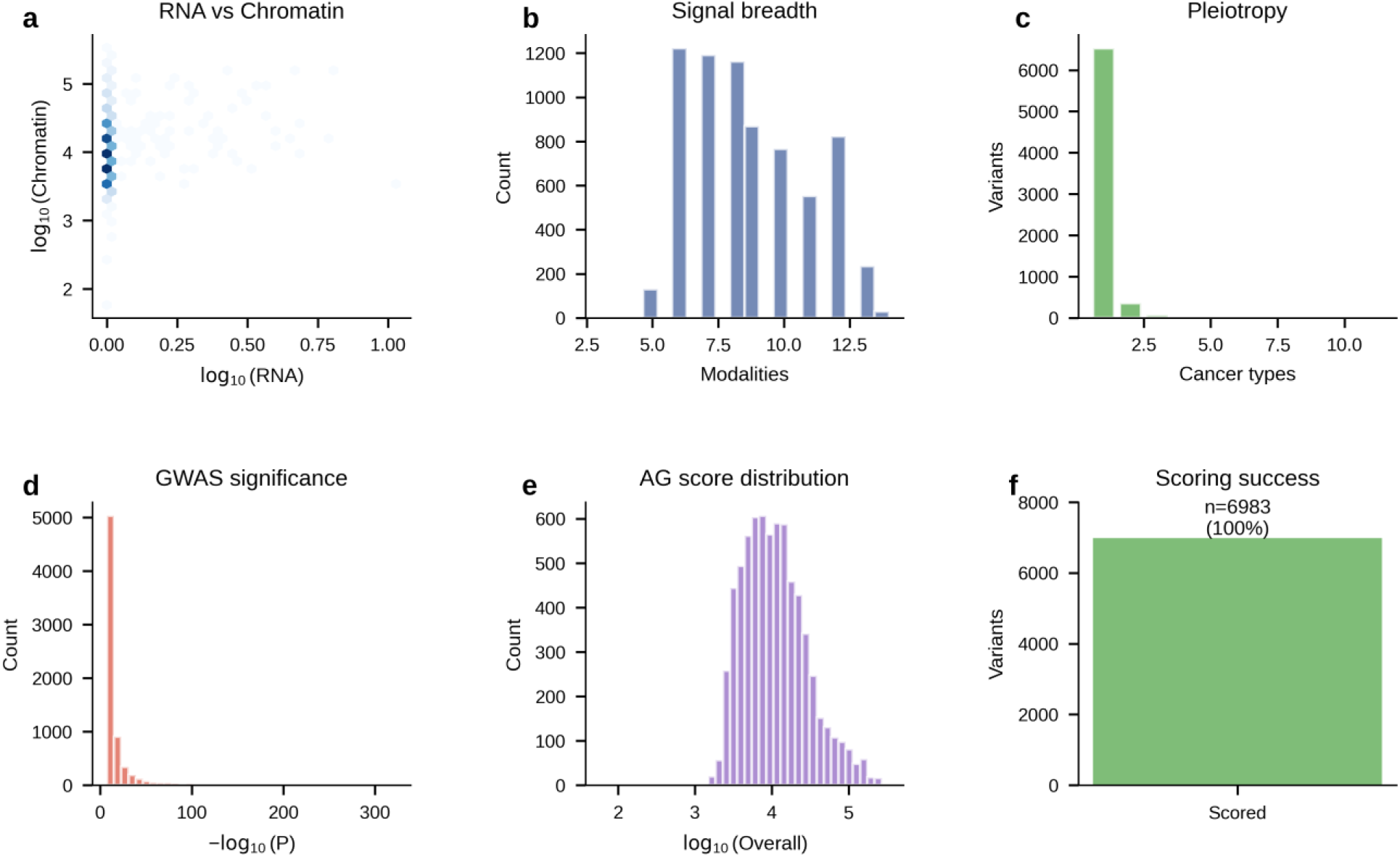
Data quality and variant characteristics. (a) RNA versus chromatin AlphaGenome effects. (b) Distribution of multimodal signal breadth. (c) Distribution of pleiotropy across cancer types. (d) Distribution of GWAS association significance. (e) AlphaGenome overall score distribution. (f) AlphaGenome scoring success across atlas variants.

**Figure S2.**
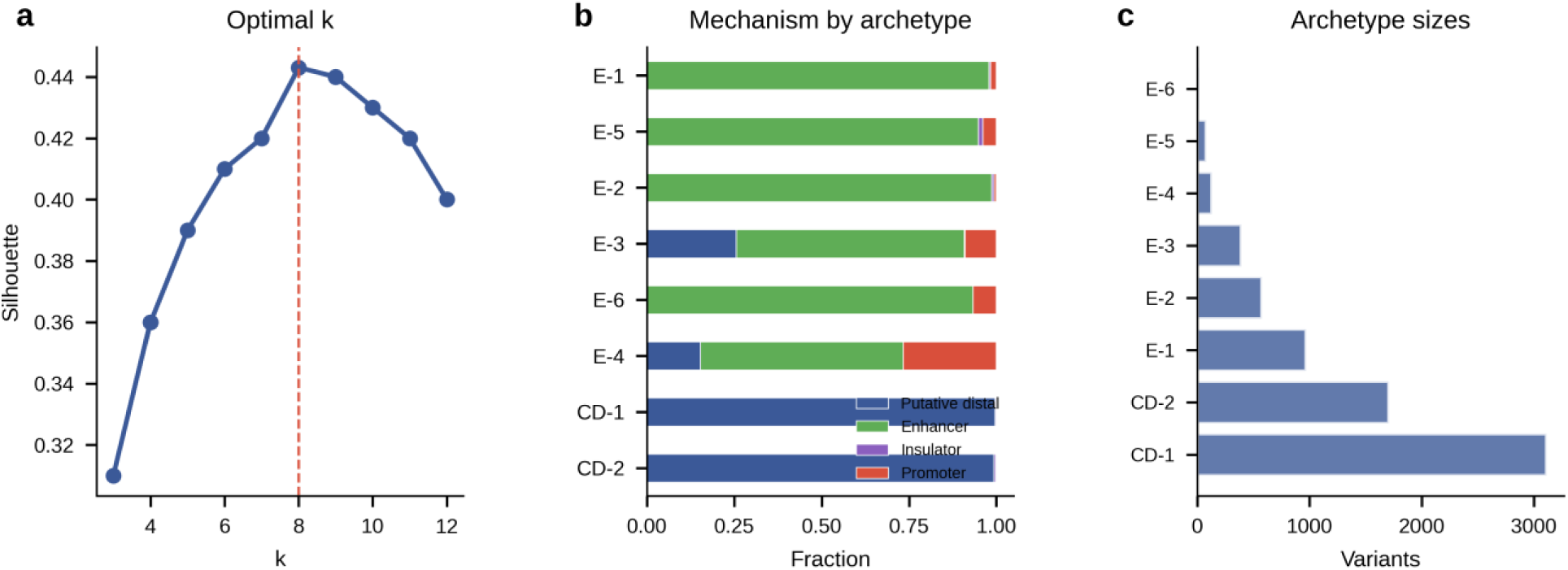
Extended archetype analysis. (a) Silhouette score by k (optimal k = 8). (b) Mechanism class composition per archetype (stacked bar). (c) Archetype sizes.

**Figure S3.**
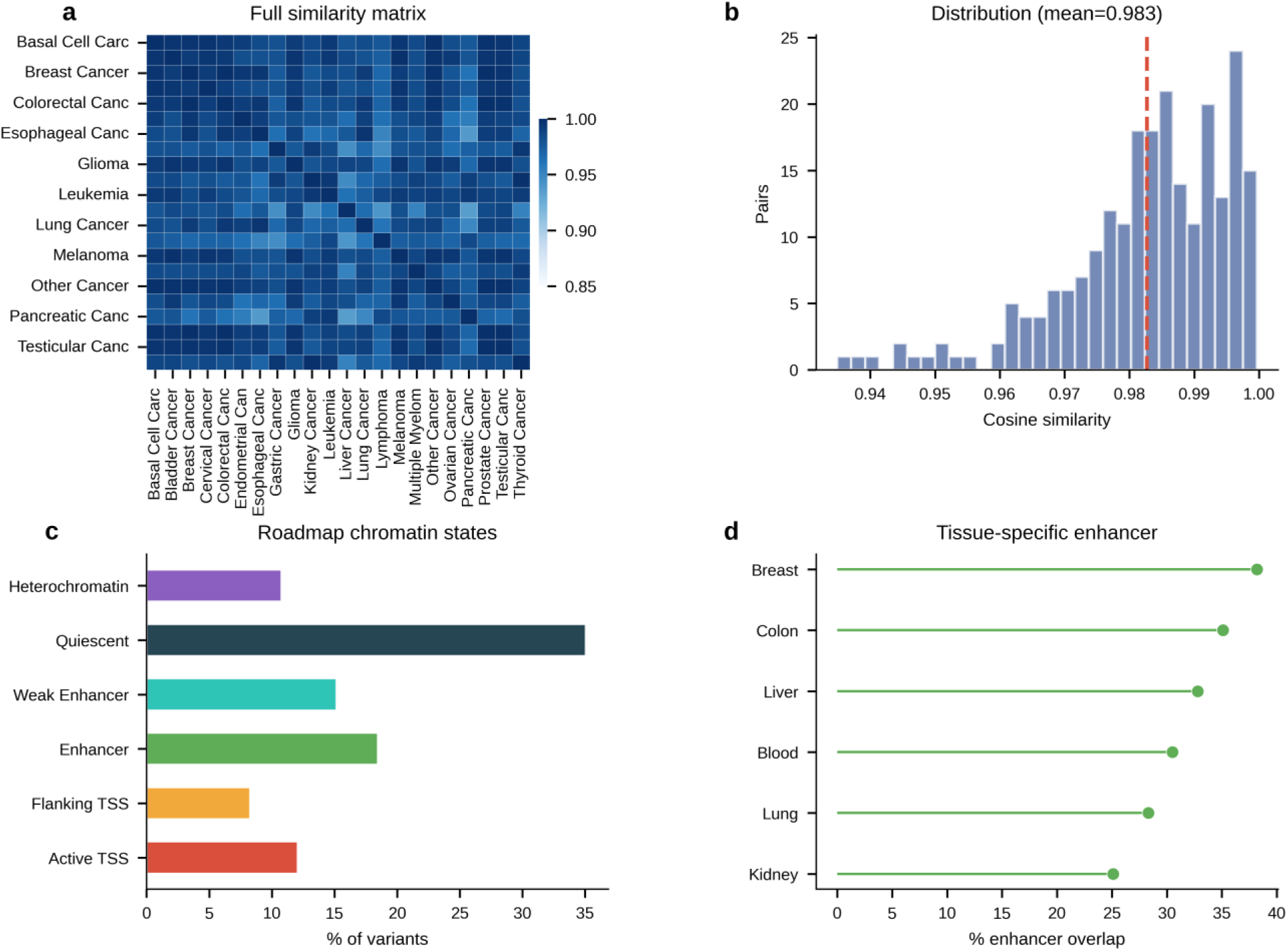
Pan-cancer similarity and tissue enrichment. (a) Full cancer-type regulatory similarity matrix. (b) Distribution of pairwise cosine similarity values across cancer-type pairs. (c) Roadmap Epigenomics chromatin-state overlap across atlas variants. (d) Tissue-specific enhancer overlap across selected Roadmap epigenomes.

**Figure S4.**
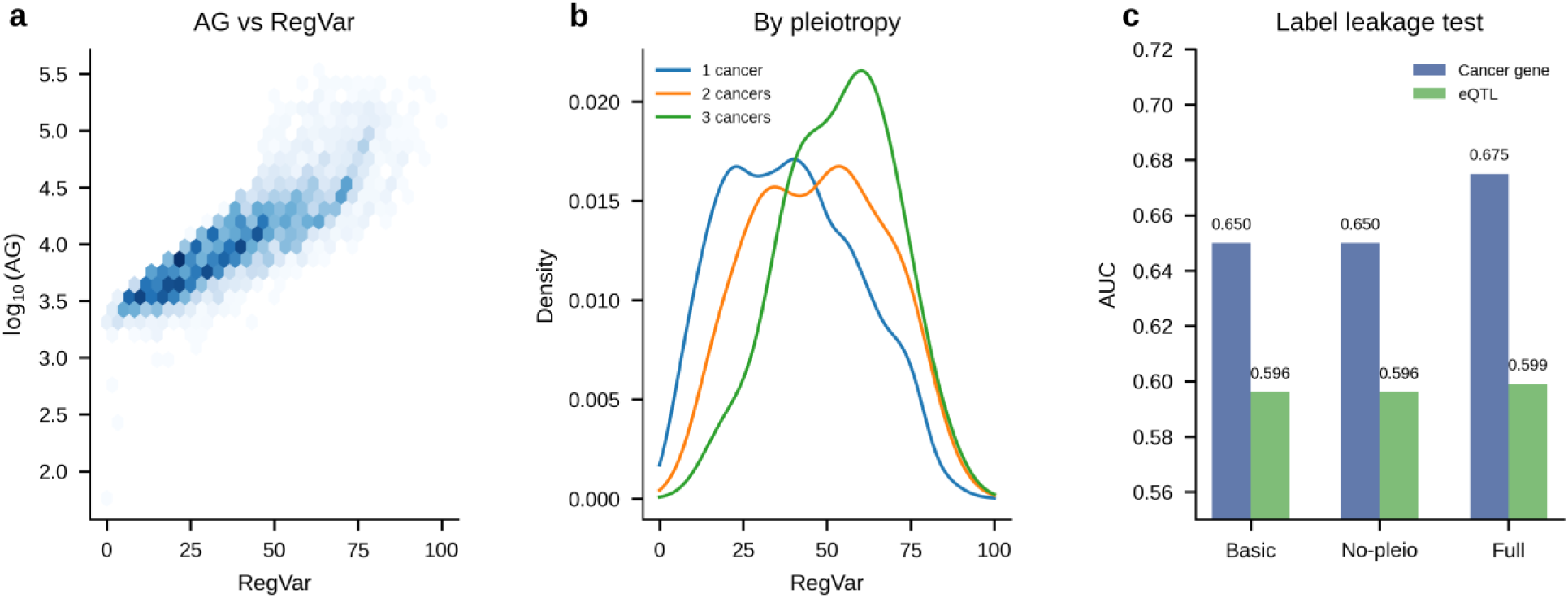
RegVar component and version analysis. (a) AlphaGenome versus RegVar (hexbin). (b) RegVar score distributions stratified by pleiotropy level. (c) Three RegVar versions: basic AUC = 0.650, no-pleiotropy 0.650, full 0.675 (demonstrates label-leakage-free performance).

**Figure S5.**
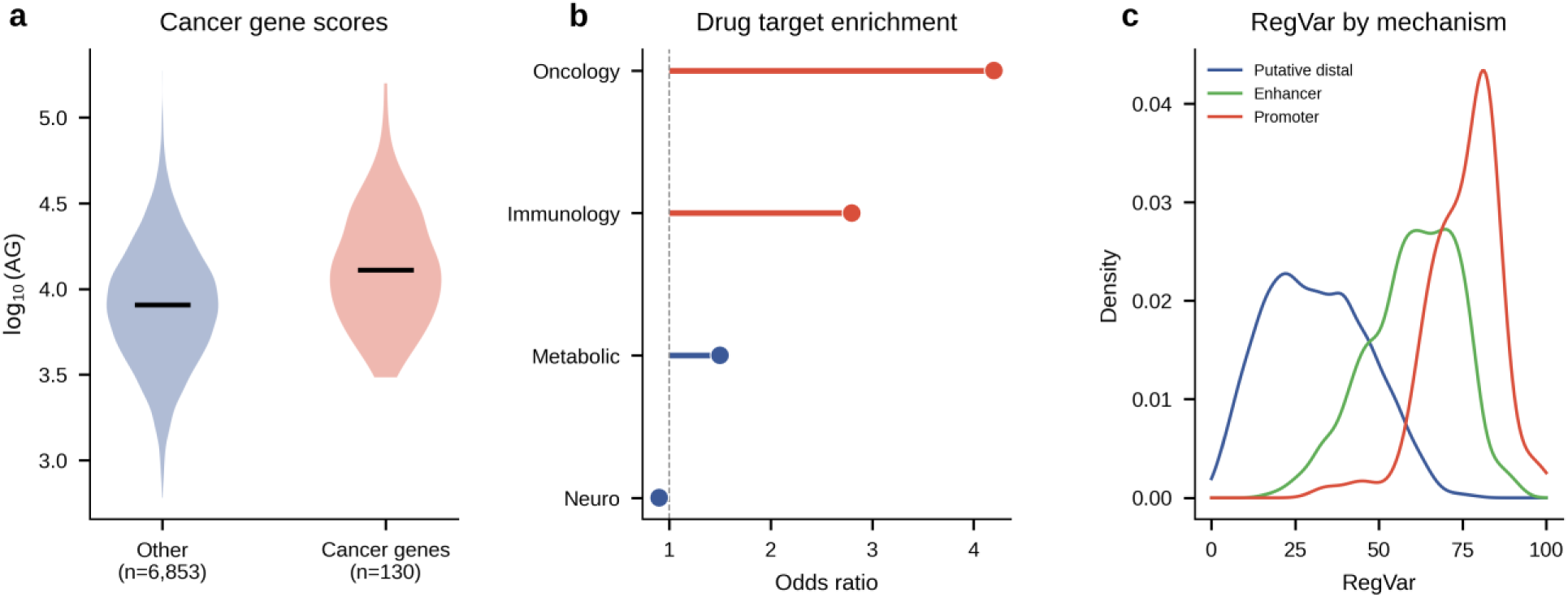
Extended validation. (a) AlphaGenome scores for cancer gene targets versus other genes. (b) Drug target enrichment by therapeutic category. (c) RegVar score distributions stratified by mechanism class.

**Figure S6.**
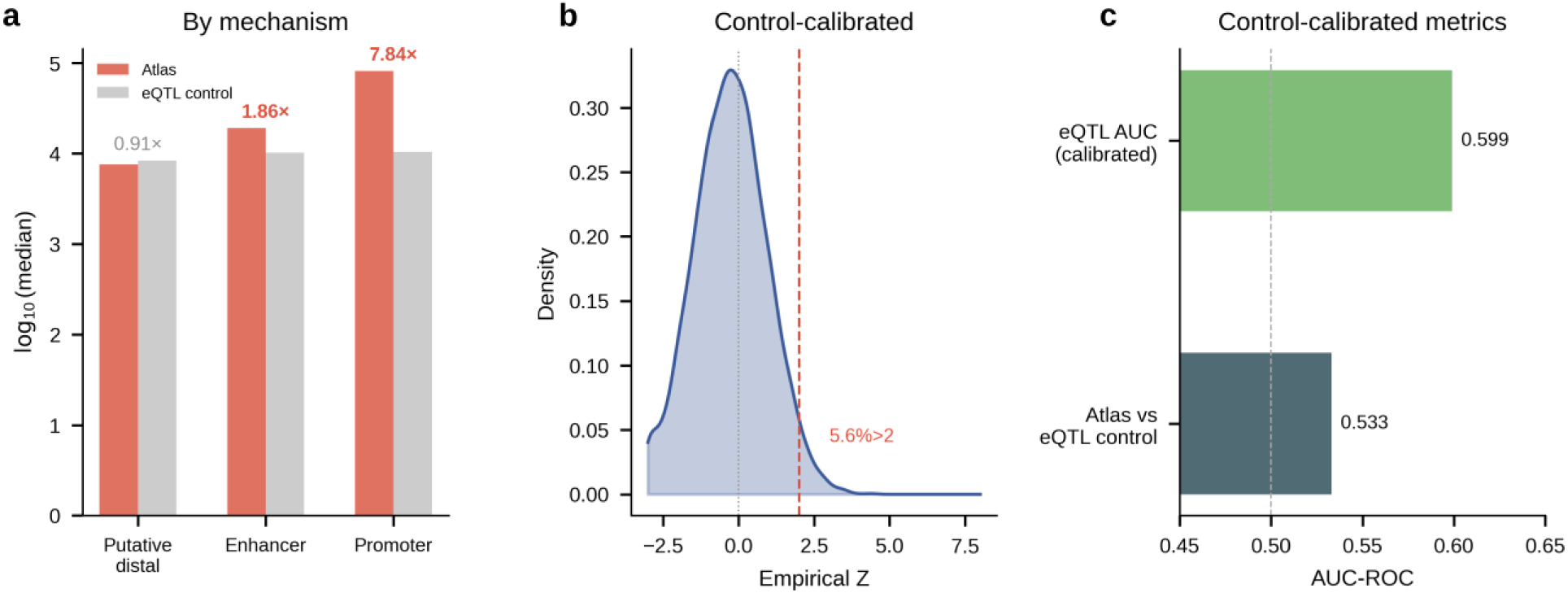
Position-matched eQTL control variant comparison. (a) Median AlphaGenome scores for atlas versus position-matched eQTL control variants by mechanism class; enhancer 1.86×, promoter 7.84×, putative distal 0.91×. (b) Empirical Z-score distribution; 5.6% of variants had Z > 2. (c) Control-calibrated AUC metrics for eQTL prediction and atlas-versus-control discrimination.

**Figure S7.**
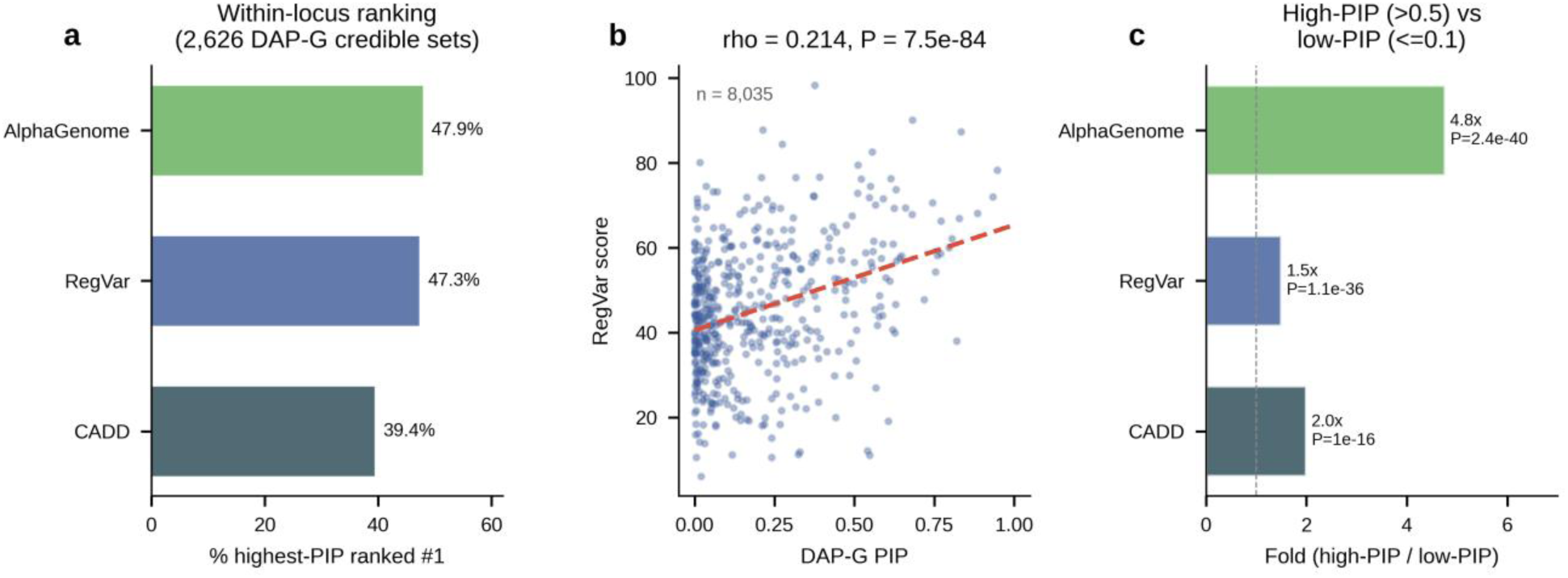
Within-locus fine-mapping validation using GTEx DAP-G eQTL credible sets. (a) Percentage of 2,626 credible sets where the highest-PIP variant is ranked #1 by each score (RegVar 47.3%, AlphaGenome 47.9%, CADD 39.4%; mean percentile ranks shown). (b) PIP-RegVar correlation across 8,035 variant-locus observations (rho = 0.214, P = 7.5 × 10^−^⁸⁴). (c) Fold difference between high-PIP (>0.5) and low-PIP (≤0.1) variant median scores.

**Figure S8.**
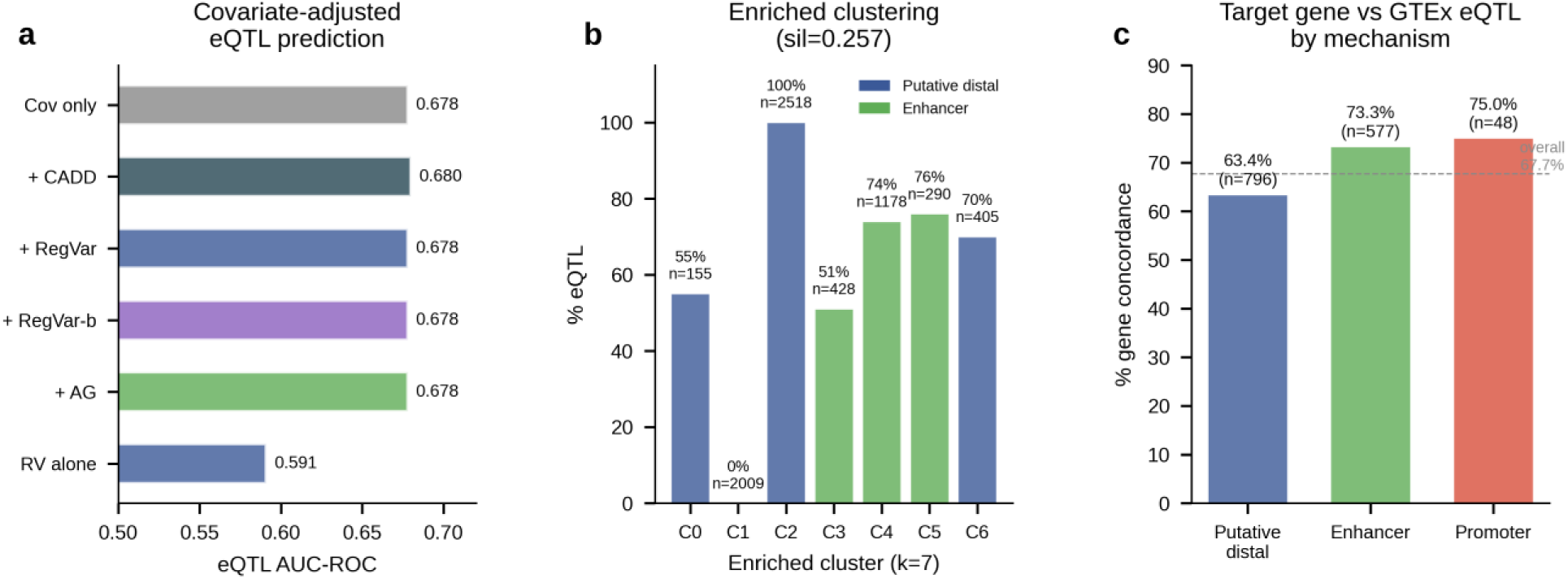
Covariate-adjusted analysis, enriched clustering, and target-gene concordance by mechanism. (a) Logistic regression AUC for eQTL prediction: covariates alone (TSS distance + cCRE class + gene density) achieve AUC = 0.678, with minimal incremental contribution from any score. (b) Enriched clustering (k = 7, 8 features) showing eQTL rate per cluster; one cluster is 100% eQTL (n = 2,518), another 0% (n = 2,009). (c) Target-gene concordance with GTEx eQTL by mechanism class: promoter 75.0%, enhancer 73.3%, putative distal 63.4%.

### Supplementary Note 1: Motif Analysis and Permutation Controls

Raw PWM disruption counts showed several high-count motifs (NR2C2, HOXA7, VAX2) among the most disrupted across the full atlas. In the 2,000-variant permutation subset used for calibration, the observed counts for these motifs (Table S4) did not exceed dinucleotide-shuffled background, indicating that these counts are explained by motif base composition rather than true enrichment (NR2C2 Z=−0.43, HOXA7 Z=0.90, VAX2 Z=−0.13, all NS). The permutation-enriched candidate transcription factors span immune signaling (NFKB1/NF-κB Z=6.67, STAT1 Z=4.97, IRF1 Z=2.24) and environmental/chromatin response (ARNT Z=5.14, HIC2 Z=4.67, ZNF354C Z=5.52). These results underscore that statistical calibration is essential for interpreting motif disruption frequencies.

### Supplementary Note 2: Archetype Robustness

The primary clustering used 4 quantitative features + mechanism indicators (k=8, silhouette=0.443). As a robustness check, enriched clustering with 8 features (adding CADD, pleiotropy, TF disruptions, TSS distance, eQTL status) yielded k=7, silhouette=0.257. The enriched clustering produced biologically interpretable separation: one cluster was 100% eQTL (n=2,518), another 0% eQTL (n=2,009), and others showed intermediate rates (51–76%). The lower silhouette reflects the higher dimensionality; the eQTL-based descriptive separation illustrates how inclusion of functional annotations stratifies the variant set, but should not be interpreted as independent validation because eQTL status was included as an input feature.

### Supplementary Note 3: RegVar Weight Optimization

Grid search over 282 weight combinations with 5-fold CV. Optimized weights: AG=0.50, mechanism=0.15, TF=0.05, breadth=0.10, pleiotropy=0.20 (RegVar-full AUC=0.675 vs RegVar-basic AUC=0.650, difference=0.025). All top-5 configurations shared increased AG weight and decreased TF weight, indicating that AlphaGenome effect magnitude is the strongest single predictor. Three-version analysis (Table S2) confirms that removing GWAS-derived features reduces AUC by only 0.025, demonstrating that the score is not inflated by label leakage.

### Supplementary Note 4: Multi-ancestry Considerations

Of 6,983 variants, only 20 have documented multi-ancestry replication, including rs6983267 (EUR+EAS+AFR), rs10069690 (EUR+AFR), and rs3803662 (EUR+EAS+AFR). AlphaGenome scores did not differ between multi-ancestry and EUR-only variants (P=0.15), though this comparison is underpowered. Extension to multi-ancestry GWAS through GBMI and All of Us will be needed for generalizability.

### Supplementary Note 5: Covariate-Adjusted eQTL Analysis

Logistic regression showed that covariates alone (TSS distance + cCRE class + gene density) predict eQTL status with AUC=0.678. Adding any score provided minimal incremental AUC (≤0.002). This indicates that the raw eQTL AUC differences (RegVar-full 0.599 vs CADD 0.488) partly reflect the association between scores and known genomic features (proximity to genes, cCRE overlap). The primary utility of RegVar and AlphaGenome lies in within-locus ranking of candidate regulatory variants in eQTL credible sets (Table S5), where they significantly outperform random while CADD does not.

### Supplementary Methods

#### GWAS Catalog Data Processing

Cancer-associated variants were extracted from the NHGRI-EBI GWAS Catalog (release r2026-04-27, 1,099,366 total associations) using the gwas_catalog Python package. Disease traits were mapped to 23 cancer categories using Experimental Factor Ontology (EFO) terms: breast carcinoma (EFO:0000305), prostate carcinoma (EFO:0001663), colorectal cancer (EFO:0005842), lung carcinoma (EFO:0001071), melanoma (EFO:0000756), basal cell carcinoma (EFO:0004593), bladder carcinoma (EFO:0000292), renal cell carcinoma (EFO:0000681), thyroid carcinoma (EFO:0002892), ovarian carcinoma (EFO:0001075), and 13 additional cancer types. Treatment-response, pharmacogenomic, survival, and toxicity associations were excluded by filtering on EFO parent terms. Variants were lifted to GRCh38 coordinates where necessary using the UCSC liftOver tool. Duplicate entries at identical genomic positions were collapsed, retaining the most significant P-value and concatenating cancer type annotations with pipe delimiters. Independent loci were defined by merging variants within 500-kb windows on the same chromosome, yielding 1,302 loci. This window-based definition was chosen as a conservative approximation in the absence of LD reference panel data for all 23 cancer types.

#### AlphaGenome Scoring Pipeline

Variant effect prediction used the AlphaGenome Python SDK (v1.0) via the public API. For each variant, a 1,048,576-bp input sequence centered on the variant position was extracted from the GRCh38 reference genome (GCA_000001405.15_GRCh38_no_alt_analysis_set.fna). The AlphaGenome model returns predictions across 4,887 output tracks spanning 19 functional modalities. Four summary statistics were extracted per variant using a custom feature extraction function: (1) maximum absolute RNA-seq effect across 667 cell types, (2) maximum absolute chromatin effect across all accessibility and histone tracks, (3) maximum absolute effect across all modalities (overall effect), and (4) number of modalities with detectable signal (defined as any track exceeding 2 standard deviations from the variant-centered baseline). For 1,678 variants (24.0%) where the GWAS Catalog did not specify a risk allele, all three possible alternate alleles were scored and the allele producing the maximum absolute overall effect was retained. Scoring was performed on the HUMMEL-2 HPC cluster with checkpointing every 25 variants to enable resumption. Total scoring time was approximately 20 hours for the full atlas.

#### ENCODE-4 cCRE Integration

Candidate cis-regulatory elements (cCREs) were obtained from the ENCODE-4 registry (ENCODE Project Consortium, 2020; 1,063,878 human elements on GRCh38). Each atlas variant was classified by direct positional overlap with cCRE boundaries: PLS (promoter-like signature, n = 98 variants, 1.4%), pELS (proximal enhancer-like signature) or dELS (distal enhancer-like signature, combined n = 1,935, 27.7%), CTCF-only (insulator, n = 28, 0.4%), or no cCRE overlap (putative distal regulatory, n = 4,922, 70.5%). Overlap was assessed using bedtools intersect (v2.30) with a minimum 1-bp overlap requirement.

#### RegVar Score Construction

RegVar integrates five component scores into a weighted composite: (1) AlphaGenome effect magnitude, computed as the percentile rank of the maximum overall effect across all atlas variants (range 0–100); (2) mechanism class score (promoter = 100, enhancer = 75, insulator = 50, putative distal = 25); (3) TF motif disruption bonus (100 if ≥1 JASPAR PWM disruption detected, 0 otherwise); (4) multimodal signal breadth, computed as the percentile rank of the number of modalities with signal (range 0–100); and (5) cross-cancer pleiotropy score, computed as (n_cancers – 1) × 20, capped at 100. Weights were optimized by exhaustive grid search over 282 combinations (increments of 0.05 per component, constrained to sum to 1.0) with 5-fold cross-validated AUC-ROC against the cancer gene discrimination task. Optimized weights: AlphaGenome = 0.50, mechanism = 0.15, TF disruption = 0.05, breadth = 0.10, pleiotropy = 0.20. Cross-validation folds were randomly assigned at the variant level; locus-aware stratification was not applied but would be preferable for future iterations.

#### Position-Matched eQTL Control Variant Scoring

The control variant pool comprised 1,277,232 unique genomic positions from GTEx v8 Whole Blood significant eQTL variant-gene pairs (FDR < 0.05). For each of the 6,983 atlas variants, one control variant was selected from the same chromosome within ±500 kb, excluding positions already in the atlas. Selection was randomized with a fixed seed (numpy random seed 42) for reproducibility. A total of 6,626 position-matched eQTL control variants were successfully identified (357 atlas variants lacked eligible control candidates within the distance window). All control variants were scored using the identical AlphaGenome pipeline: same SDK version, same 1,048,576-bp window, same max-of-3-alternate-alleles strategy, and same feature extraction code. The control pool consists of eQTL-positive variants, making the comparison conservative; additionally, the ±500-kb proximity constraint means control variants may share partial LD with atlas variants, further attenuating observed differences.

#### Within-Locus Fine-Mapping Validation

GTEx v8 DAP-G eQTL fine-mapping results were downloaded from the GTEx Portal (GTEx_v8_finemapping_DAPG.CS95.txt.gz, 43 MB compressed). This file contains 1,447,425 variant entries across 418,575 eQTL credible sets (95% coverage) in 49 tissues. Each entry encodes variant-cluster membership as ENSG_ID:cluster_number@Tissue=PIP pipe-delimited strings in column 6. For the within-locus ranking test, credible sets containing two or more atlas variants were identified (n = 2,626 clusters, comprising 8,035 variant-locus observations). Within each cluster, atlas variants were ranked by RegVar, AlphaGenome, and CADD scores, and the percentile position of the highest-PIP variant was recorded (0 = top-ranked, 1 = bottom-ranked). Mean percentile rank was tested against the null expectation of 0.5 using a one-sample Wilcoxon signed-rank test. PIP-score correlations were assessed by Spearman rank correlation.

